# Modelling COVID-19 in the North American region with a metapopulation network and Kalman filter

**DOI:** 10.1101/2024.06.05.24308495

**Authors:** Matteo Perini, Teresa K. Yamana, Marta Galanti, Jiyeon Suh, Roselyn Kaondera-Shava, Jeffrey Shaman

## Abstract

**Background:** Metapopulation models provide platforms for understanding infectious disease dynamics and predicting clinical outcomes across interconnected populations, particularly for large epidemics and pandemics like COVID-19.

**Methods:** We developed a novel metapopulation model for simulating respiratory virus transmission in the North America region, specifically for the 96 states, provinces, and territories of Canada, Mexico and the United States. The model is informed by COVID-19 case data, which are assimilated using the Ensemble Adjustment Kalman filter (EAKF), a Bayesian inference algorithm, and commuting and mobility data, which are used to build and adjust the network and movement across locations on a daily basis.

**Findings:** This model-inference system provides estimates of transmission dynamics, infection rates, and ascertainment rates for each of the 96 locations from January 2020 to March 2021. The results highlight differences in disease dynamics and ascertainment among the three countries.

**Interpretation:** The metapopulation structure enables rapid simulation at large scale, and the data assimilation method makes the system responsive to changes in system dynamics. This model can serve as a versatile platform for modeling other infectious diseases across the North American region.

**Funding:** US Centers for Disease Control and Prevention Contract 75D30122C14289; US NIH Grant AI163023.

## Introduction

Mathematical models have been used to simulate infectious diseases outcomes, infer transmission dynamics, and predict future disease burden. These tools can inform public health strategies by testing control methods and identifying effective interventions^1^. During the SARS-CoV-2 pandemic, unprecedented data availability enabled application of mathematical models in many locations and at different geographical scales worldwide^2–5^. Metapopulation modeling approaches provide an efficient framework for simulating and evaluating the spatiotemporal progression of infectious disease over large geographic areas. In this model form, populations are aggregated within typically fixed geographic units (e.g. provinces, cities), which allows resolution of spatial disease patterns without the computational expense and micro-behavioral assumptions required for agent-based models. At large geographical scales, agent-based models require high-performance computing (HPC) clusters. For example, Bhattacharya et al. developed a platform to enable the real-time execution of an agent-based COVID-19 model for the United States on more than ten thousand CPU cores^6^.

Many metapopulation models have been developed to describe the dynamics of infectious diseases at different geographic scales^7,8^; however, a key challenge in model development lies in accurately determining the movement patterns of individuals among subpopulations. Some metapopulation models use fixed or arbitrary sized geographical areas (cells) as subpopulations, which can then be aggregated to match the resolution of available case, census and movement data^9,10^. One example of a multi-national system is the GLEAM (Global Epidemic And Mobility) platform^10–12^, which estimates the flux of individuals among arbitrary subpopulations centered around major transportation hubs (usually airports) and uses commuting and air travel data^13^.

In 2020, the US reported the highest number of COVID-19 cases and deaths globally, with the first case identified in Washington state on January 20^th^ and three pandemic waves manifesting during the year^14^. Canada reported its first case in Toronto on January 25^th^ and experienced a decline in cases during the summer followed by a resurgence in the fall. Mexico had a similar epidemiological history, but the first wave developed later during the summer^15^. The literature currently lacks a comprehensive COVID-19 model for the North American region at continental scale. Here, we present a metapopulation susceptible-exposed-infectious-recovered (SEIR) for COVID-19 for the majority of the North American region (i.e. Canada, United States and Mexico) from the beginning of the pandemic to widespread availability and distribution COVID-19 vaccines. The model is coupled with a data assimilation algorithm and is informed by daily COVID-19 cases and a commuting network that is adjusted by daily mobility trends. Unlike the GLEAM platform, in this model the flux of individuals among subpopulations of the metapopulation model is calculated using the daily work commuting patterns coupled with random movements among the geographical locations, as in previous work^4^.

The model developed here provides temporal estimates of transmission and ascertainment rates for COVID-19 for the North American Region, filling a gap in the existing literature regarding models for this region. The flux patterns and the continental-level metapopulation structure used in this study helps investigation of disease dynamics of COVID-19 at a larger scale, for the majority of the North American region, and can reveal dynamics that are discernible only at such a broad scale. Lastly, the work commuting matrix structure developed for this study can be paired to different compartmental models and disease surveillance data to study the dynamics of other infectious diseases such as influenza.

## Methods

We developed a metapopulation susceptible-exposed-infectious-recovered (SEIR) model for the North American region. Specifically, the model represents the 96 first-level administrative divisions of Canada (10 provinces and 3 territories), United States (50 states and 1 federal district) and Mexico (31 states and 1 autonomous city) represented in **Figure 1**, with a total population of 483 million, based on census data^16–18^. Mixing is simulated as two types of movement: daily commuting and random movement (i.e. all the daily movement across divisions due to reasons other than work commuting).

**Figure 1.**
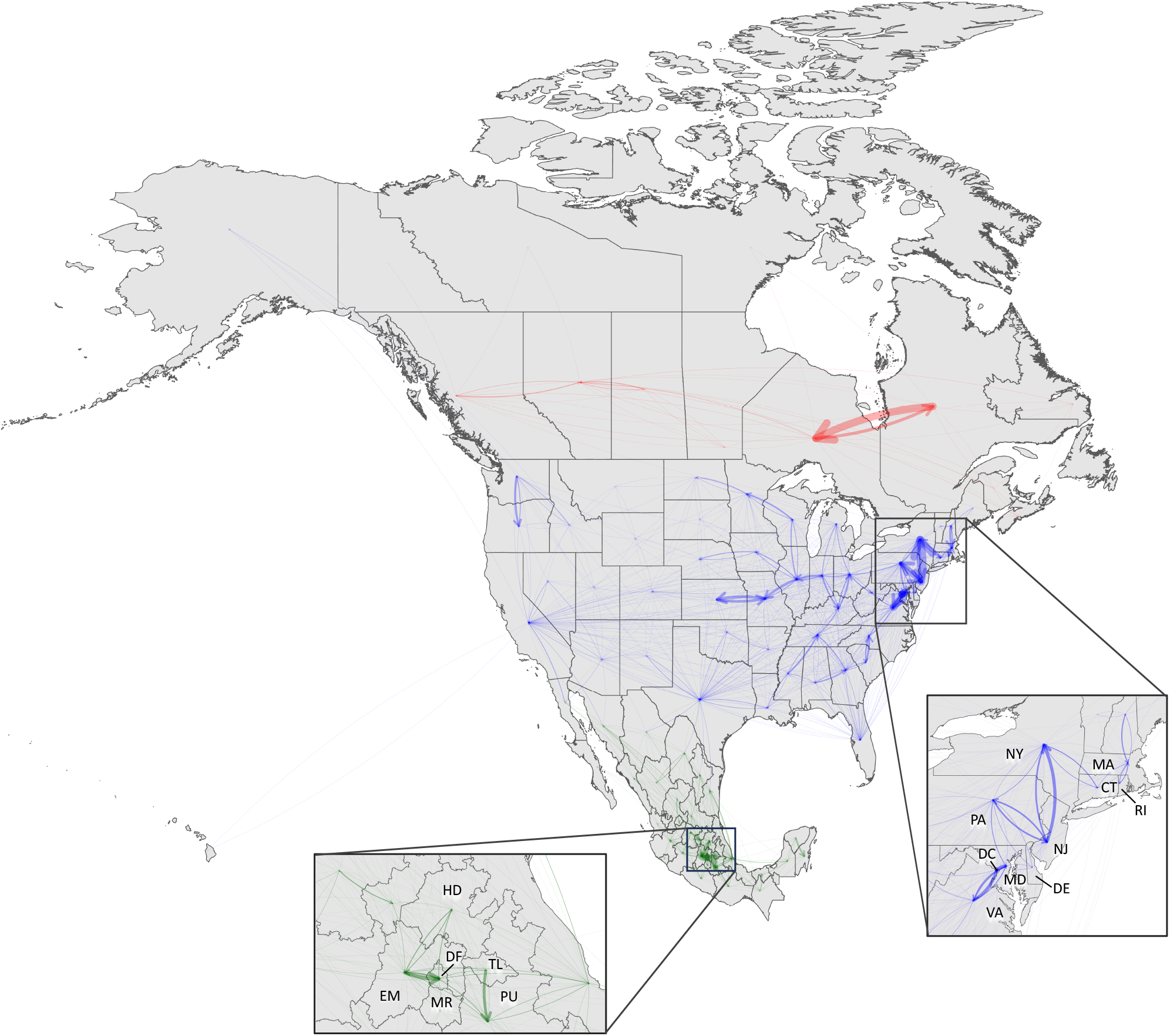
Commuting to work matrix in most of the North American region. The map shows the 10 provinces and 3 territories of Canada, the 50 states and 1 federal district of United States and the 31 states and 1 autonomous city of Mexico that have been used in the model. The arrows represent the flux of individuals commuting daily to work to another location. Arrow size represents the number of commuters, and color represents the country of origin: red for Canada, blue for United States, green for Mexico.

### Daily commuting matrix

Daily commuting among locations were retrieved and derived from four national datasets: i) Canadian 2016 census (Statistics Canada) Commuting Flow from Geography of Residence to Geography of Work^19^; ii) Canada Frontier Counts (Statistics Canada): Number of vehicles travelling between Canada and the United States^20^; iii) 2011-2015 5-Year American Community Survey (ACS) Commuting Flows (United States Census Bureau)^21^; and iv) Mexican Intercensal Survey 2015 (National Institute of Statistics and Geography, INEGI)^18^. Information from these datasets were processed and combined according to the methods described in **Supplementary Note 1** to obtain the commuting work matrix represented in **Figure 1**. The matrix contains the number of people that commute daily to work in another location (states for Mexico and US, provinces or territories for Canada). To account for control measures and closures enacted during the estimation period, the commuting work matrix was scaled based on daily mobility activity, derived from the “change in workplace visitors” trends from Google Community Mobility Reports^22^ (see **Supplementary Figure 1**). To account for movement between locations for purposes other than work commuting, the model includes daily random movement among locations; this movement was set to be proportional to the average number of working commuters among each location pair (see **Table 1**).

**Table 1:**
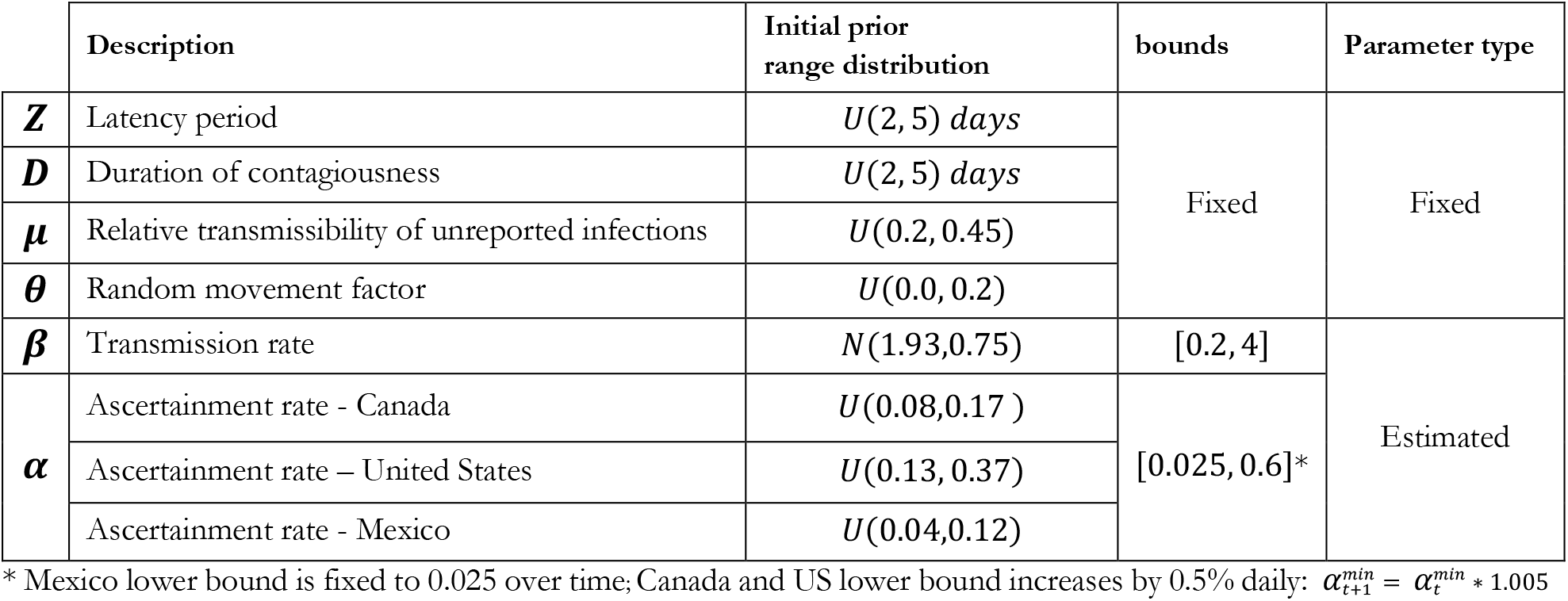
parameters description, initial prior distribution, ranges, and parameter type.

### Transmission model

The metapopulation model resolves daytime and nighttime mixing differences, depicting diurnal changes in contact among subpopulations. Transmission occurs as a discrete Markov process during both day and nighttime, following the structure of previous studies in the US^4^. The transmission dynamics are described by the equations in **Supplementary Note 2** (**eqs. S4**-**S13**). In these equations, 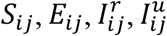, and *N*_*ij*_ represent the susceptible, exposed, reported infectious, unreported infectious, and total population in the subpopulation commuting from location *j* to location *i* (*i ← j*). Additionally, we assume that no individuals enter or leave the model, and that there is no loss of immunity following primary infection, given the relatively short simulation time period. For the same reason, we can compute the *R*_*ij*_ (Removed population) as 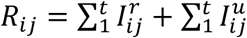 which is the cumulative sum of daily new reported and unreported cases, or the sum of the individuals that exited the two infectious compartments.

The parameters of the model are: *β*, the transmission rate of reported infections; *µ*, the relative transmissibility of unreported infections; *Z*, the average latency period (from infection to contagiousness); *D*, as the average duration of contagiousness; *α*, the fraction of documented infections (ascertainment rate); and *θ*, a multiplicative factor adjusting random movement. A distinct transmission rate, *µβ*, is defined for undocumented infections: we assume that these individuals show little to no symptoms during infection and are less contagious than documented infections^5^. Each equation is integrated using a Poisson process to capture the stochastic nature of transmission dynamics. In total, the model consists of 3,268 metapopulations. To reduce the dimension of the system being estimated and to improve identifiability, we fixed the values of the parameters related to disease progression (*Z, D*, and *µ*) and the multiplicative factor for random movement (*θ*) based on previously published findings^4,14^. Local transmission rates 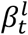 and ascertainment rates 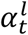 are estimated for each of the 96 locations, *l* at time *t*. The hyperparameter 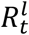, the time-varying reproductive number, was derived using the next-generation matrix approach^4,23^:

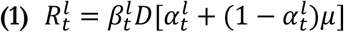

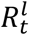 Time-varying reproductive number for location *l* at time *t*

*D* Duration of contagiousness

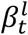 Transmission rate of location *l* at time *t*

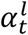 ascertainment rate of location *l* at time *t*

*µ* relative transmissibility of unreported infections

### COVID-19 cases and data assimilation

The model is informed by daily COVID-19 confirmed case data retrieved from COVID-19 Open Data — Google Health^24^ starting from January 20^th^ 2020 to March 31^st^ 2021 for the 96 first-level administrative divisions of Canada, United States and Mexico. This time frame was chosen to capture the first three COVID-19 waves^14^, before the emergence of the Delta and subsequent variants and more widespread uptake of COVID-19 vaccines. A 7-day moving average was applied to the daily cases data to smooth out daily fluctuations in reporting. The smoothing also mitigates the impact of reporting delays or inconsistencies in case data, providing a more reliable indicator of overall disease dynamics.

The estimation of state variables and parameters is carried out by data assimilation implemented using the Ensemble Adjustment Kalman Filter (EAKF) algorithm^25^, as in previous studies^26,27^. Kalman filters use Bayes’ rule to update state variables and parameters. Normality is assumed for the prior distribution and the likelihood so that the posterior distribution can be characterized by the mean and the covariance. In this model, an ensemble of 300 simulations is integrated to generate a prior distribution of parameters and state variables, including estimation of the observed state variable. At each observation time point, the model is halted, and the ensemble and observation are used to calculate the Kalman gain, which is used to update the observed state variable. The unobserved state variables and parameters are then updated in proportion to the same Kalman gain. Finally, the posterior estimates are used as priors and model integration through time continues to the next observation. The formulas used to calculate the Kalman gain for the observed and unobserved variables and parameters are presented in the **Supplementary Note 3** (**eqs, S14**-**S15**). To properly balance the influence of the observational data in the assimilation process, it is crucial to estimate its error, which is typically unknown. Here, we estimated the Observational Error Variance of the observational data as shown in **eq. (2)**, similarly to prior works^5,28,29^:

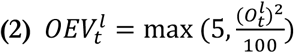

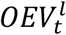 Observational Error Variance of location *l* at time *t*

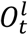 average cases in location *l* in the week before time *t*

Repeated filter adjustments tend to decrease model ensemble variance reducing the impact of new observations in subsequent estimates. This may lead to divergence, in which the filter ceases adjusting the model state^25^. To avoid divergence, at each timestep we applied a multiplicative factor (1.01) to inflate the prior ensemble of the observed variable (daily reported infected *I*^*r*^) and the estimated parameters (ascertainment rate *α* and transmission rate *β*). Additionally, we reinitialized the values of the estimated parameters for a fraction (2%) of the ensemble members every 7 days. This reinitialization enables the system to periodically readjust estimations whenever the ensemble variance begins to shrink, thus preventing divergence.

Further details on the SEIR-EAKF assimilation process are available in the **Supplementary Note 3**.

### Model initialization

Each ensemble member was initialized with a set of parameter and state variable estimates to resemble the epidemiological conditions at the beginning of the SARS-CoV-2 pandemic in 2020. The parameter values related to disease progression (*Z, D* and *µ*) were drawn from distributions with ranges reported in **Table 1**, in accordance with Li et al^5^. These parameters are assumed to remain fixed over time and to have consistent values across all locations, representing inherent biological characteristics of the disease. The random movement factor *θ* is also constant over time, with a random value drawn from a uniform distribution between 0 and 0.2 assigned to each location (see **Table 1**). This value represents the relative volume of random movement compared to commuting, where *θ* = 0.15 indicates that the number of random visitors is 15% of the average number of commuters between two locations. Exposed (*E*), reported infectious (*I*^*r*^) and unreported infectious (*I*^*u*^) individuals were initialized with random draws from a uniform distribution that ranged from 1 to 9 in each subpopulation.

The three countries exhibited substantive differences in healthcare and testing capacity during the pandemic. Mexico had particularly low testing rates^30,31^, suggesting that only severe cases were assayed due to limited availability of test kits. During 2020, the United States experienced the highest number of cases, as well as relatively high nation-wide ascertainment rates, as shown in prior inference studies^4,14^. For Canada, the estimate of national testing rates or the ascertainment rate is lacking; however, Ontario, the most populous province in Canada, accounting for ∼37% of the population, initially faced challenges in ramping up its testing capacity, but its centralized resource strategy was able to increase test capacity during 2020^32^. Given these differences among the three countries, we assigned country-specific initial ranges for the ascertainment rate *α*. For the US we used SARS-CoV-2 seroprevalence estimates and cumulative reported cases to estimate the value of the ascertainment rate. Specifically we used infection-induced seroprevalence from blood samples collected in US during July 2020 (3.5%)^33^ to derive an initial estimate of *α* in the US, this formula is shown in **eq. (3)**:

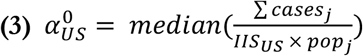

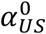 Initial ascertainment rate for the US

∑ *cases*_*j*_ Cumulative reported cases of state *j* up to July 2020

*IIS*_*US*_ Infection-induced seroprevalence in the United States during July 2020 (3.5%)^33^

*pop*_*j*_ Population of state *j*

Based on this formula, we set the initial prior distribution mean to 0.25 for *α* in US.

Infection-induced seroprevalence was not available for Canada and Mexico during the time frame of interest. Instead, considering the very low testing rate for Mexico^30,31^ we set the initial prior distribution mean to 0.08, corresponding to a ∼70% decrease from the US. Canada faced early issues establishing testing facilities^32^, so we set the initial prior distribution mean to 0.12 for *α*, between the values of US and Mexico (∼50% decrease compared to the US).

The initial values of *β* were drawn for each location from the normal distribution with mean *µ* = 1.93 and standard deviation *σ* = 0.75 similarly to Li et al.^5^(see **Table 1**).

The SEIR-EAKF model-inference system can potentially estimate parameters and state variables values that violate physicality. This could happen, for example, if a state variable or estimated parameter is adjusted by the filter to a value below zero. To avoid this issue, we applied constraints to both the state variables and parameters, subsequently re-assigning values that fell outside specified bounds. Specifically, ensemble members associated with any state variable that exhibited values less than or equal to zero were assigned values from the previous day. The Susceptible population *S*_!”_ is not directly adjusted by the EAKF. Instead, it is computed using the total population count and the EAKF-adjusted state variables as *S*_*ij*_ = *N*−*E*_*ij*_ −*R*_*ij*_. This guarantees population mass balance and is feasible under the assumption of non-reinfection among individuals. Additionally, it prevents the variable *S*_*ij*_ from exceeding the total population, which aligns with the assumption of a constant population.

The lower bound of *α* was set to 0.025 corresponding to 5 reported cases every 200 infections. For Canada and US, this bound increased linearly by 0.5% at each day, or: 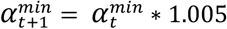. This increase was imposed to reflect the efforts of the two countries to increase the detection capabilities of local and national health systems^32,34–36^. Conversely, Mexico implemented a sentinel surveillance system^37^ in which only the hospitalized cases and 10% of the mild cases were tested^38^. Focusing testing efforts on symptomatic individuals is a cost-effective strategy, but it also greatly increases the proportion of unreported cases, resulting in lower ascertainment rates. For this reason, the lower *α* bound of Mexico did not increase over time in our model. The upper bound for *α* was fixed for all locations at 0.6 (i.e. 60 cases per 100 infections). We allowed the transmission rate *β* to span a broad range of values, with the lower bound set to 0.2 and the upper bound set to 4. The descriptions, prior ranges and bounds for all the model parameters are shown in **Table 1**.

### System identifiability

To assess the identifiability of the model, we first tested our framework using synthetically generated datasets. Specifically, we generated a suite of synthetic outbreaks using the model (**eqs. S4-S13**) in free simulation, each with arbitrarily assigned values of the epidemiological parameters and initial conditions for state variables. We then ran the full model-inference system 100 times assimilating the daily new reported cases time series generated by each of these free simulations to test the system’s ability to accurately estimate state variable and parameter values. As in practice with actual data, we fixed the parameters related to disease progression (*Z, D*, and *µ*) and the multiplicative factor for random movement (*θ*), while the ascertainment rate *α* and transmission rate *β* were estimated for each location. The initial prior distribution and range of the parameters were as reported in **Table 1**.

## Results

### Daily commuting matrix

The daily commuting matrix depicted in **Figure 1** was obtained by combining and processing the national datasets reported in the Method section (full description in **Supplementary Note 1**); it is summarized in **Table 2**. Out of a combined population of 483 million, the Canadian population accounts for the 7.3% (35.1 million), the US for the 67.2% (324.3 million) and Mexico for the 25.6% (123.3 million). Around 8.9 million people, or 1.84% of the represented North American population, commutes daily to another state/province/territory or country to work. Inter-country commuting accounts for just the 1.12% (99,369) of the 8.9 million commuters. Finally, the percentage of internal commuters for each country is similar to the population percentage relative to the total population of the three countries.

**Table 2:** Number of commuters by country of residence and workplace. Individuals with residence and workplace in the same country are commuting between states/provinces/territories within that country. The last column shows the percentage over the total commuters. The last row shows the total number of cross-border (state, province, territory, or country) commuters and its percentage over the total population of the model.

| Residence | Workplace | Commuters | % over Total Commuters |
| --- | --- | --- | --- |
| Canada | Canada | 527,300 | 5.93 |
| Canada | USA | 1,962 | 0.02 |
| USA | Canada | 4,605 | 0.05 |
| USA | USA | 5,334,072 | 60.02 |
| USA | Mexico | 12,892 | 0.15 |
| Mexico | USA | 79,910 | 0.90 |
| Mexico | Mexico | 2,925,820 | 32.92 |
|  |  | **Total Commuters** | **% over total Population** |
|  |  | 8,890,144 | 1.84 |

### System Identifiability

To verify the convergence of the estimated parameters (ascertainment rate *α* and transmission rate *β*) to the synthetic truth values created in free simulation, we plotted the model-generated data points over the boxplot distributions of the 3000 estimated parameter values (300 ensemble members x 100 ensemble simulations) at the end of each outbreak among the locations (see **Supplementary Figure 2**). Most of the true values fall in the interquartile of the distribution of the estimated values (68% for *α* and 84% for *β*) while all the true values falls in the 95% CI of the distribution. This demonstrates the ability of the system to estimate local time-varying *α* and *β* values.

**Figure 2.**
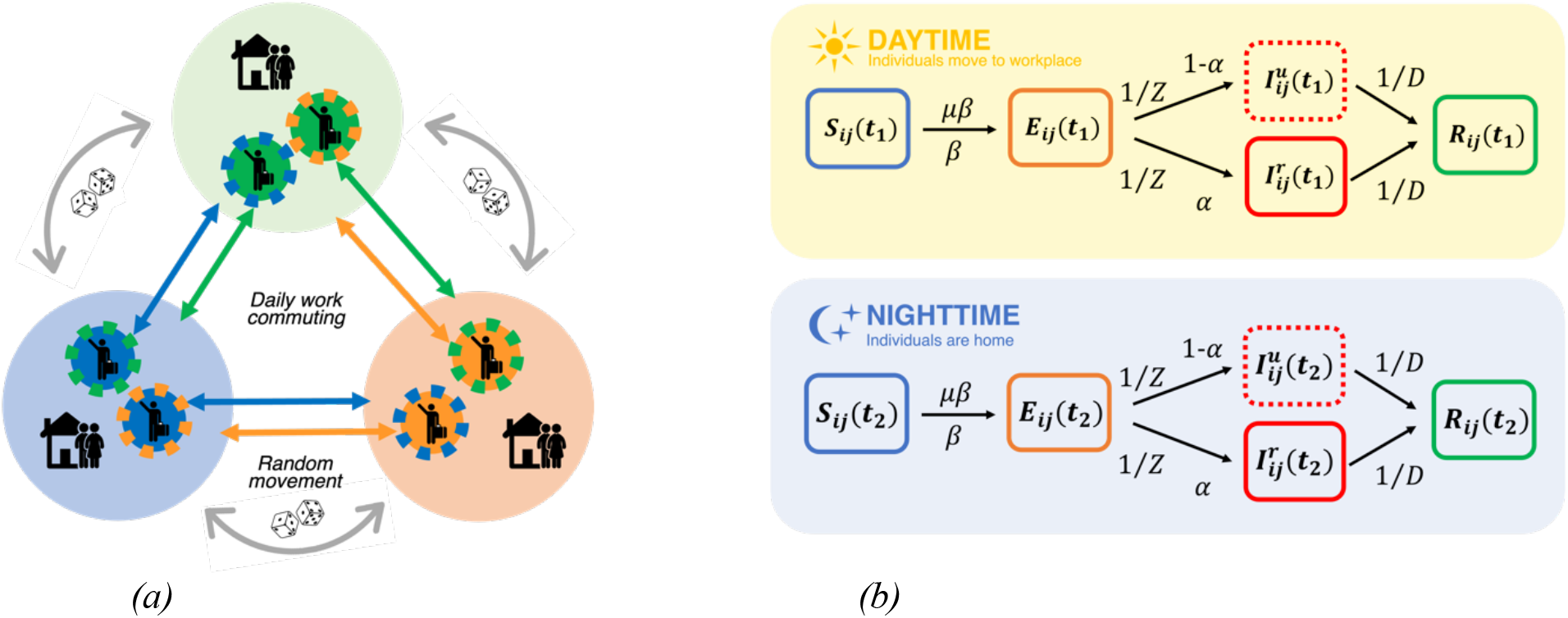
Metapopulation structure and compartmental model: **(a)** Daily work commuting - during the daytime, some individuals commute from their home to their workplace in another location and mix with the populations present there. During the nighttime, those commuters return home and mix with other residents who live in the same location. Random movement - individuals may travel among locations for reasons other than work. These random visitors circulate among subpopulations following a Markov process, causing a population exchange in all locations **(b)** 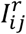 reported infected and 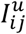 unreported infected from location from location j to location i (i ←j); t_1_ daytime duration; t_2_ nighttime duration; β transmission rate; *α* ascertainment rate; µ relative transmissibility of unreported cases; Z latency period; D duration of contagiousness.

### Simulation with Case Data

The model-inference system was run with the real case data of Canada, United States and Mexico starting from January 20^th^ 2020 to March 31^st^ 2021. During this time frame, the three countries experienced asynchronous outbreaks as depicted in **Supplementary Figure 1**. To explore disease dynamics over time and location, we selected three timepoints when cases were declining in most of the locations of the North American region, roughly corresponding to the end of the three pandemic waves experienced in United States during 2020^14^. The first wave began in January 2020 and lasted through the spring; we selected June 6, 2020 as the first timepoint. **Figure 3** shows the estimated values of the parameters (ascertainment rate *α* and transmission rate *β*) and hyperparameters (basic reproductive number *R*_*t*_) in all 96 locations of the study region at this time point. In addition, the lower panels of **Figure 3-5** show the time progression of model fitting (modeled daily new reported cases and observed daily cases), select state variables (*S*, cumulative *I*^*r*^,cumulative *I*^*u*^) and parameter and hyperparameter estimates (*α*, *β, R*_*t*_) for a total of nine selected locations. These locations were selected among the 96 to represent the epidemiological progression in different geographical areas of the North American region, focusing on some of the most populous and epidemiologically relevant locations of the three countries. In **Figure 3** the three selected locations indicated in the maps are British Columbia (Canada), New York (United States) and Distrito Federal (Mexico City, Mexico). The sub-national parameter values for each of the three countries were aggregated to the national level using a population-weighted average. On June 6, 2020, the population-weighted average ascertainment rate *α* of the mean estimates in US states (0.26) was 1.5 times higher than in Canada (0.17) and 2.7 times higher than in Mexico (0.10). The population-weighted average transmission rate *β* of the mean estimates for Mexican states was 1.03, which is 1.4 times higher than in Canada (0.72) and the US (0.72). The population-weighted average of the time-varying reproductive number *R*_*t*_ of the mean estimates was 0.99 for Canadian provinces and territories, 1.11 for the United States, and 1.22 for Mexican states. These values are also reported in **Table 3**.

**Table 3:** Population-weighted average of the mean estimates values of the parameters (ascertainment rate *α* and transmission rate β) and hyperparameters (time-varying basic reproductive number R_t_) for the three countries at three selected timepoints. The intensity of cell colors in the table corresponds to their values: higher values are represented by more intense colors.

| Location | Jun. 6<sup>th</sup> 2020 | Sep. 7<sup>th</sup> 2020 | Mar. 15<sup>th</sup> 2021 |
| --- | --- | --- | --- |
| **Ascertainment rate $\alpha$** |  |  |  |
| Canada | 0.17 | 0.18 | 0.26 |
| United States | 0.26 | 0.31 | 0.36 |
| Mexico | 0.10 | 0.11 | 0.13 |
| **Transmission rate $\beta$** |  |  |  |
| Canada | 0.72 | 0.95 | 0.84 |
| United States | 0.72 | 0.64 | 0.85 |
| Mexico | 1.03 | 0.86 | 0.85 |
| **Time-varying basic reproduction number $R_t$** |  |  |  |
| Canada | 0.99 | 1.32 | 1.31 |
| United States | 1.11 | 1.04 | 1.49 |
| Mexico | 1.22 | 1.04 | 1.07 |

**Figure 3.**
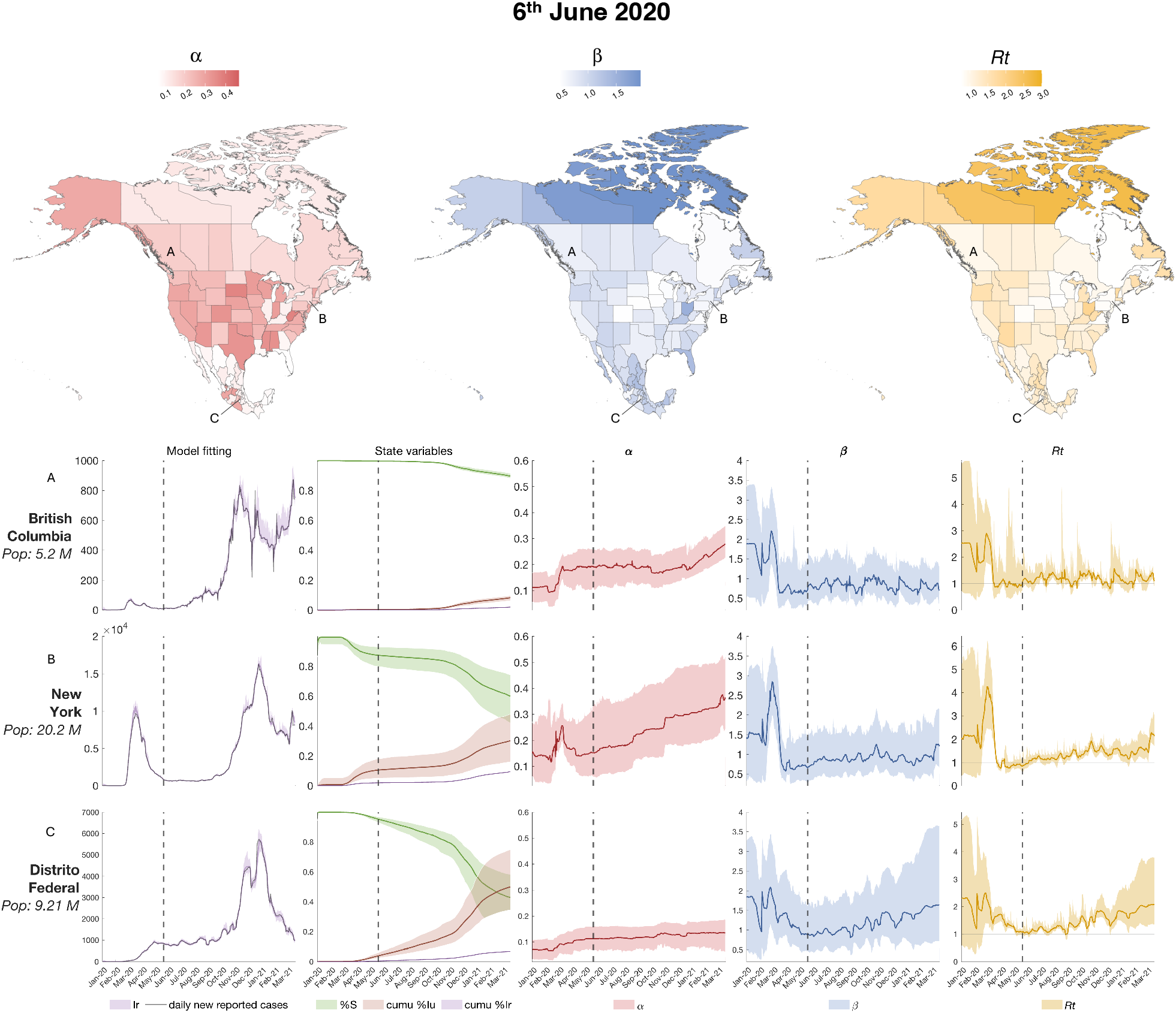
Estimated state variables and parameters on June 6, 2020: the three maps on the top panel show the value of the of the parameters (ascertainment rate *α* and transmission rate β) and the hyperparameter (basic reproductive number R_t_) for all 96 locations on June 6, 2020. The bottom panels show the model fitting (i.e. the estimated observed variable over the 7-day smoothed daily new reported cases), three state variables illustrating disease progression (the susceptible population, cumulative reported infectious and cumulative unreported infectious), and the parameters α, β and R_t_ for 3 selected locations: British Columbia (Canada), New York (US) and Distrito Federal (Mexico City, Mexico). The color shaded areas represent the 95% credible interval from the 300-member ensemble. The dotted vertical lines indicate the timepoint of reference for the maps.

During summer 2020, some locations, particularly in the US, experienced a second wave consisting of a resurgence of cases, with a decline at the beginning of fall. **Figure 4** shows the values of the state variables and parameters on September 7, 2020. The maps show the parameter estimates across all locations, while the temporal dynamics of state variable and parameter estimates are shown for three of the nine selected locations: Ontario (Canada), Florida (US) and Estado de México (Mexico). On September 7, 2020, the population-weighted average ascertainment rate *α* of the mean estimates in the United States was 0.31, which is 1.74 times higher than in Canada (0.17) and 2.8 times higher than in Mexico (0.11). The population-weighted average transmission rate *β* of the mean estimates in Canadian provinces and territories was 0.95, which is 1.5 times higher than in the US (0.64) and 1.1 times higher than in Mexican (0.86). The population-weighted average of the time-varying reproductive number *R*_*t*_ of the mean estimates was 1.32 for Canadian provinces and territories, 1.26 times higher than in the United States (1.04) and Mexican states (1.04). These values are also reported in **Table 3**.

**Figure 4.**
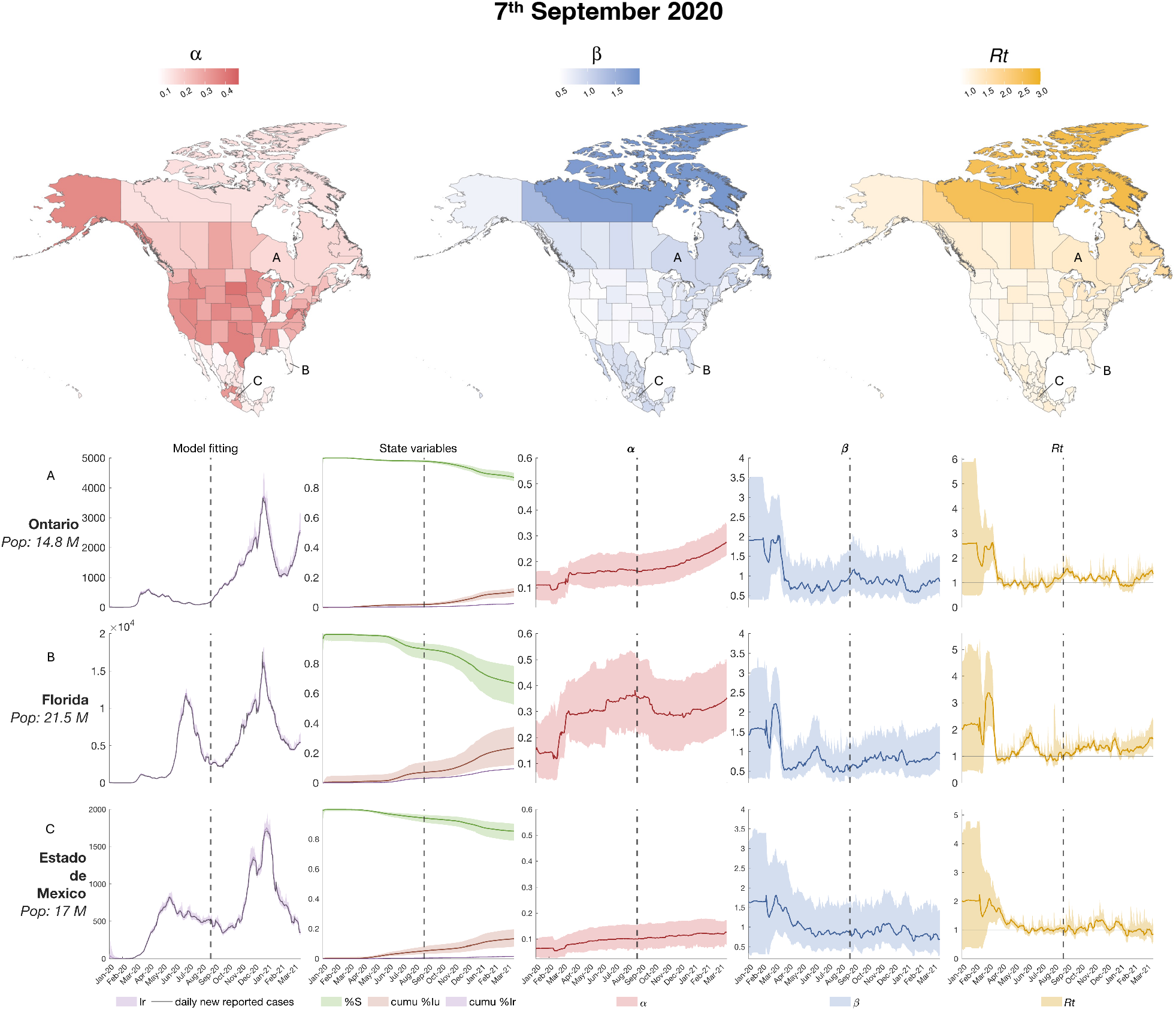
Estimated state variables and parameters on September 7, 2020: the three maps on the top panel show the value of the of the parameters (ascertainment rate *α* and transmission rate β) and the hyperparameter (basic reproductive number R_t_) for all 96 locations on September 7, 2020. The bottom panels show the model fitting (i.e. the estimated observed variable over the 7-day smoothed daily new reported cases), three state variables illustrating disease progression (the susceptible population, cumulative reported infectious and cumulative unreported infectious), and the parameters *α*, β and R_!_ for 3 selected locations: Ontario (Canada), Florida (US) and Estado de México (Mexico). The color shaded areas represent the 95% credible interval from the 300-member ensemble. The dotted vertical lines indicate the timepoint of reference for the maps.

Most locations experienced a more severe outbreak during the autumn-winter wave of 2020/2021, before the widespread availability of the COVID-19 vaccines. We plot the estimated parameters and hyperparameters on March 15, 2021 in **Figure 5**. The state variable and parameter estimate time series are also shown for three of the nine selected locations: Quebec (Canada), California (United States), and Jalisco (Mexico). On March 15, 2021, the population-weighted average ascertainment rate *α* of the mean estimates in the United States was 0.36, 1.4 times higher than Canada (0.26) and 2.7 times more than Mexico (0.13). The population-weighted average transmission rate *β* of the mean estimates was 0.83 in Canada and 0.85 in US and Mexico. The population-weighted average of the time-varying reproductive number *R*_%_ was 1.49 for the United States, 1.14 times higher than Canadian provinces and territories (1.31) and 1.39 times higher than Mexican states (1.04). These values are also reported in **Table 3**. All the estimated mean values of the parameters (ascertainment rate *α* and transmission rate *β*) and hyperparameters (basic reproductive number *R*_%_) for all the location on these three timepoints are reported in **Supplementary Table 3**.

**Figure 5.**
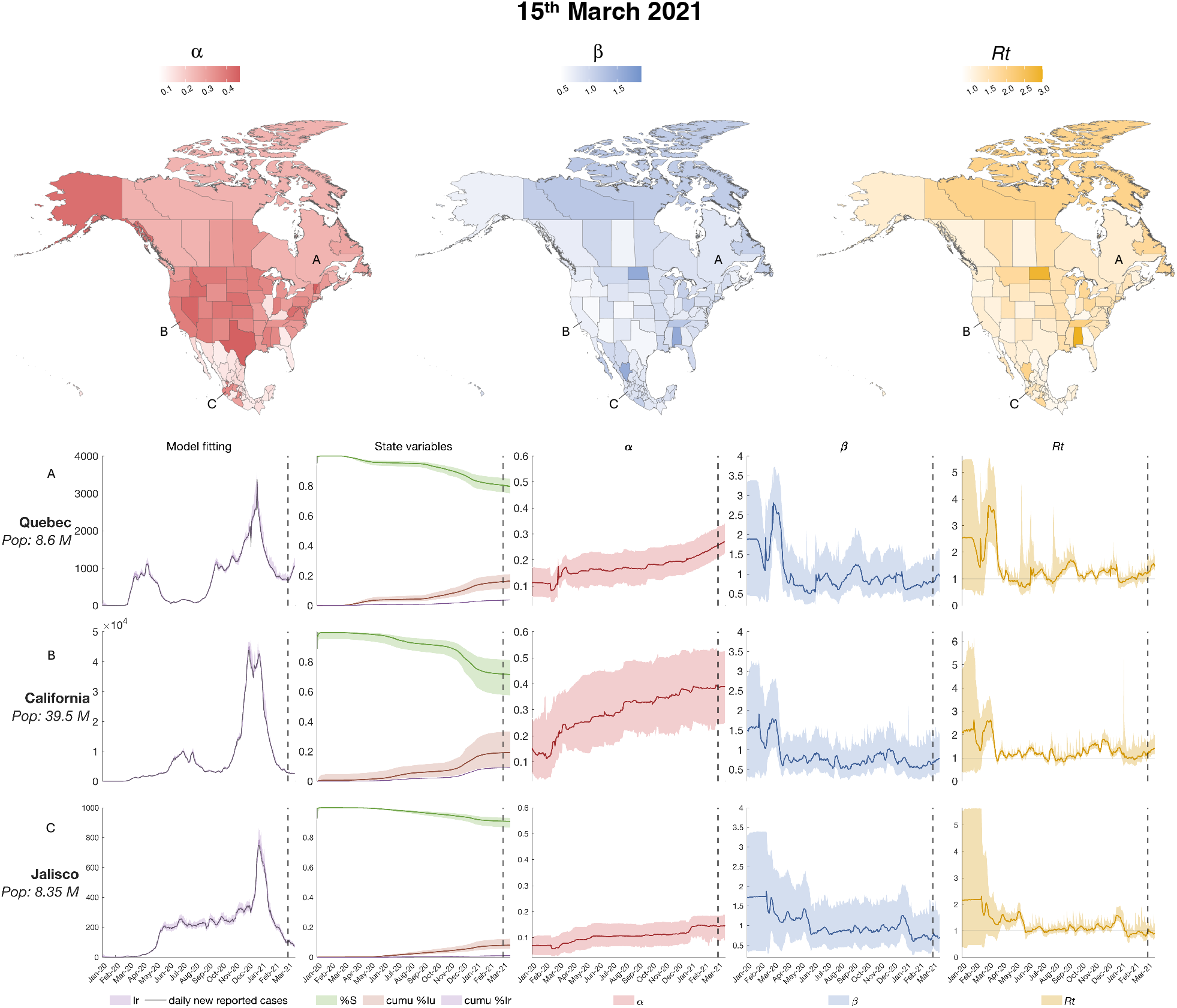
Estimated state variables and parameters on March 15, 2021: the three maps on the top panel show the value of the of the parameters (ascertainment rate *α* and transmission rate β) and the hyperparameter (basic reproductive number R_t_) for all 96 locations on March 15, 2021. The bottom panels show the model fitting (i.e. the estimated observed variable over the 7-day smoothed daily new reported cases), three state variables illustrating disease progression (the susceptible population, cumulative reported infectious and cumulative unreported infectious), and the parameters *α*, β and R_t_ for 3 selected locations: Quebec (Canada), California (United States), and Jalisco (Mexico). The color shaded areas represent the 95% credible interval from the 300-member ensemble. The dotted vertical lines indicate the timepoint of reference for the maps.

## Discussion

In this study we developed a SEIR metapopulation model structure and combined it with a data assimilation algorithm (EAKF) to reproduce COVID-19 outbreak dynamics and estimate important epidemiological parameters across most of the North American region during the first three pandemic waves^14^. The model has demonstrated identifiability in estimating system state variables, as well as the ascertainment rates, *α*, and transmission rates, *β*, for the 96 first-level administrative divisions of Canada, United States, and Mexico. The metapopulation structure provided computational efficiency in comparison to agent based models that require HPC (high-performance computing) and cluster computing approaches to run at large scale^6^. The transmission module of the model structure relies on the daily work commuting patterns across states, provinces, and territories for disease spreading, with the capability to adjust the daily commuting matrix using Google Mobility Report trends data^22^ to account for travel restrictions implemented during the pandemic.

As depicted for selected locations in **Figure 3-5** and reported for all the locations in **Supplementary Table 3**, the SEIR-EAKF system estimated large disparities in the ascertainment rates *α* among the three countries^4,14,30–32^. Moreover, it estimated a gradual increase in the ascertainment rates *α* throughout the three major COVID-19 outbreaks during 2020 and at the beginning of 2021 in each country. Specifically, the US showed substantially higher *α* values in almost all states across the three waves: at the last time point (March 15, 2021) the majority of states had reached an ascertainment rate of ∼40%. Canada and Mexico showed smaller ascertainment rates than the US at the end of the estimation: most of the Canadian provinces and territories had an ascertainment rate of ∼26%, while Mexican states had less than 20%. Note that in Canada and the United States, the increase in *α* is partially influenced by the increasing lower imposed during inference, whereas Mexico has a fixed *α* lower bound over time.

Transmission rates *β* remained generally low during the estimation period for the majority of locations. In the United States, the estimated *β* values in the majority of locations appeared to exhibit minimal variation throughout the estimation period, while in Mexico transmission rates generally decreased gradually over time. A notable exception was Distrito Federal (Mexico City), the very densely populated capital of Mexico. The relatively high *β* value at the end of the estimation in March 2021 (1.6) was not unexpected in this location. High population density allows for more contact opportunities, increasing the rate of transmission and the basic reproduction number as show in previous analysis^4^. A few additional exceptions to the national trends are discussed in the **Supplementary Note 4**.

The hyperparameter *R*_*t*_ (time-varying basic reproductive number) is proportional to *β* and *α*, as reported in **eq. (1)**. The majority of the mean estimated *R*_*t*_ values were above the epidemic threshold of *R*_*t*_ = 1 at all the three selected time points and all locations had mean *R*_*t*_ > 1 at least once among the three selected time point. Moreover, the lower estimated mean values for *R*_*t*_ never dropped below 0.75, or below 0.86 at the last time point. These values suggest a sustained epidemic in the North American region.

Overall, the three countries demonstrated distinct and quite isolated epidemiological histories. This is supported by the results presented in **Figures 3-5** and **Supplementary Table 3** where the trends estimated for the parameter and hyperparameter values can be clustered by country, highlighting the national and interconnected epidemiological developments they have undergone. This phenomenon arises from the commuting network structure depicted in **Figure 1**, where few arrows traverse national borders, illustrating three major networks (the countries) with limited interconnection.

The model developed for this work uses a new commuting matrix, based on national census data and surveys, to model worker flows in the American region (Canada, US, and Mexico). It assumes most movements are daily work commutes and accounts for additional movements with a random factor proportional to the commuting flux. Moreover, the system couples the compartmental model (SEIR) with the Ensemble Adjustment Kalman Filter (EAKF), adjusting the system state variables and parameters daily on the basis of the case data. This approach differs from other models used to estimate infectious diseases on a multi-national and/or continental scale, such as the GLEAM platfom^10–12^. Those platforms estimate the movement of individuals among various arbitrary subpopulations located around major transportation hubs and leverages commuting and air travel data. In contrast, our approach utilizes state, province, and territory boundaries to define locations and is not informed by air travel data. Instead of adjusting the estimation, the GLEAM platform uses case data and other datasets to conduct a calibration run aimed at identifying the optimal set of parameters that best fit the real data in its estimations^39^.

The role of asymptomatic individuals has been shown to be central for the spread of viruses like SARS-CoV-2 and influenza, with most transmissions occurring with exposure times exceeding an hour^40^. Therefore, it is reasonable to center a model system around work-related commuting; infectious and asymptomatic individuals share the same space with coworkers for several hours, increasing their probability of infection; subsequently, each worker returns home to their resident locations, further increasing the probability of spreading disease to their families. Conversely, the GLEAM platform focuses on global disease spread, showing that the global spatiotemporal patterns of disease spreading are mainly determined by the airline network^41^. Additionally, the inclusion of air travel data enables GLEAM to capture global phenomena such as the external introduction or reintroduction of the virus during the estimation process. In contrast, the SEIR-EAKF model developed in this work is more sensitive to local changes in trends, as it leverages case data to adjust estimations. The importance of modeling each introduction event is particularly relevant at the beginning of an epidemic. This significance decreases once the epidemic in a region is primarily driven by internal transmission dynamics. Our model accommodates the external introduction of new infections through stochastic integration instead of relying on international flight data. However, the inclusion of global air travel data can improve system estimation, particularly in the initial weeks following the introduction of virus and for epidemics less prevalent than COVID-19. These differences make the two modeling approaches particularly effective at capturing different aspects and phases of epidemics. Therefore, combining these and other approaches into a single model or multi-model ensemble could improve the capacity to estimate and predict disease parameters.

The development of multinational dynamical models entails more time and effort compared to localized models because it requires the reconciliation of heterogeneous data sources. For example, the three countries in this study conducted independent census surveys that needed to be carefully interpreted and meticulously merged to ensure the resulting multinational contact network was homogeneous. The great advantage of using realistic contact networks that encompass multiple countries is the ability to infer simultaneously across broader regions (e.g. North America), rather than comparing results from separate local inference systems, thus highlighting the geographic spread of the disease across borders. Furthermore, the inference system implemented in this study can serve as a platform for modeling other respiratory infectious diseases, such as influenza, by pairing the North American commuting network developed here with other mathematical models.

Robust models for estimating disease parameters not only help to understand disease dynamics and assess responses at different locations, but they also can be adapted for use during epidemics to generate forecasts that can help health authorities in developing and implementing more informed policies. Using spatially resolved dynamical models applied at scale, it is possible to compare how various factors such as public health policies, population density, and mobility patterns affect disease spread and control in different locations. This comparative inference helps identify the most effective strategies and conditions for controlling epidemics, providing valuable insights for tailoring public health interventions to specific regions or populations. Additionally, this model can be implemented to support monitoring systems and counterfactual simulation, enhancing public health preparedness and response by enabling data-driven and location-specific strategies that can directly improve epidemic control of the region.

## Data Availability

All data produced in the present study are available upon reasonable request to the authors and available online at https://github.com/MatteoPS/NA_SEIR-EAKF

https://github.com/MatteoPS/NA_SEIR-EAKF/blob/main/NAE_suppl_material_medRxiv_Jun5.pdf

## Contributors

Data curation: MP. Formal Analysis: MP, TKY, MG. Investigation: MP. Software: MP, TKY, MG, JS RKS. Methodology: MP, JS, RKS. Writing – original draft: MP, JS, RK. Validation: TKY, MG. Supervision: TKY, MG, JS. Writing – review & editing: TKY, MG, JS. Funding acquisition: JS. Conceptualization: JS. Project administration: JS.

## Data sharing statement

The model scripts and the input data are publicly available on GitHub https://github.com/MatteoPS/NA_SEIR-EAKF

