## Supplementary Material for "Modelling COVID-19 in the North American region with a metapopulation network and Kalman filter"

### Supplementary Note 1:

#### Detailed description of the commuting matrix

*Description of the datasets*

The commuting matrix of the North American region was generated using 4 national datasets:

1. *Canadian 2016 census (Statistics Canada): Commuting Flow from Geography of Residence to Geography of Work* https://www12.statcan.gc.ca/census-recensement/2016/dp-pd/dt-td/Rp-eng.cfm?TABID=4&LANG=E&A=W&APATH=3&DETAIL=0&DIM=0&FL=A&FREE=0&GC=0&GL=-1&GID=1355304&GK=0&GRP=1&O=D&PID=113344&PRID=10&PTYPE=109445&S=0&SHOWALL=0&SUB=0&Temporal=2017&THEME=125&VID=0&VNAMEE=&VNAMEF=&D1=0&D2=0&D3=0&D4=0&D5=0&D6=0.
   This dataset provides the average number of commuters among Canadian Census Divisions (CD). These raw data were aggregated to obtain the commuting flow at province and territory level. This dataset lacks information regarding the inter-country commuting (i.e. Canadian citizens crossing the American border to go to work).
2. *Canada Frontier Counts (Statistics Canada): Number of vehicles travelling between Canada and the United States:* https://doi.org/10.25318/2410000201-eng.
   This dataset provides a count of monthly vehicles entering Canada via land customs from 1970 to 2021. The data are divided by province/territory, mode of transportation (e.g. automobiles, trucks, other), length of stay (e.g. same day or single night) and vehicle origin (United States vehicles entering or Canadian vehicles returning). For this work, we filtered the 2015 data for United States automobiles entering and Canadian automobiles returning to each province/territory with length of stay equal to one day.
3. *2011-2015 5-Year American Community Survey (ACS) Commuting Flows (United States Census Bureau):* https://www.census.gov/data/tables/2015/demo/metro-micro/commuting-flows-2015.html
   This dataset provides the commuting flows for the United States and Puerto Rico averaged over the years 2011-2015. The dataset is provided at state and county level. The data specify for each county or state how many people commute to another county, state, or country. Estimates are obtained by pooling together the results of five years (2011-2015).
4. *Mexican Intercensal Survey 2015 (National Institute of Statistics and Geography, INEGI):* http://en.www.inegi.org.mx/programas/intercensal/2015/#Microdatos
   This dataset contains the raw results of the Mexican Intercensal Survey 2015. Presently, it is the only national survey to provide information on commuting patterns of Mexican residents. Data are derived from a sampling of roughly 10% of the population (2.9 million commuters, representing 32 million workers in Mexico as stated in a report from the "Cooperative Mobility for Competitive Megaregions" at The University of Texas at Austin https://rosap.ntl.bts.gov/view/dot/54764). The average number of daily commuters for each Mexican state was obtained by aggregating the answers from the question: "*In which State (or Country) is the business, company or place where you worked last week?*". The assumption is that anyone replying to this question would commute daily in that State/Country. The resulting counts were multiplied by 10 to scale up to include the unsampled population.

*Commuting flow among the Countries:*

We only considered the movements between bordering or "near-bordering" locations across countries (i.e. states/provinces/territories less than ~150 miles from a national border). Note, the datasets report a considerable number of people having their place of work in locations of another country that do not share a border or that is not close to their residence state; however, such distances generally make daily commuting unfeasible and they have been disregarded form the calculations of the commuting flows.

For USA and Mexico, the datasets provide the number of commuters from each state to other countries, but they do not report the destination state, i.e. they show the total number of people entering another country to work but do not specify which state within the foreign country. To distribute the commuters among the states of Mexico and USA (and vice versa) we used the formula:

(S1) $$C_{l^A \to m^B} = C_{l^A \to B} \frac{C_{m^B \to A}}{C_{B \to A}}$$

| | |
|---|---|
| $C_{l^A \to m^B}$ | commuters from state $l$ of country $A$ to state $m$ of country $B$ |
| $C_{l^A \to B}$ | commuters from state $l$ of country $A$ to all the neighboring states of country $B$ |
| $C_{m^B \to A}$ | commuters from state $m$ of country $B$ to all the neighboring states of country $A$ |
| $C_{B \to A}$ | commuters from all the neighboring states of country $B$ to country $A$ |

Using this formula, we represent the non-homogeneous nature of the passage across the US-Mexican border. All commuting occurs through the relatively few custom stations that are non-uniformly distributed among the border states of Mexico and the US.

The Canadian census does not provide information on the numbers of individuals commuting to other countries, whereas the US provides the number of commuters from each US state to Canada without specifying the province or territory of destination. To approximate the number of daily commuters from Canada to US, we combined:

- The Canadian frontier counts of vehicles crossing the border (at province/territory level to or from US)
- The US dataset of commuters to Canada (at state level to Canada).

Because people cross borders by car for reasons other than commuting, the number of vehicles crossing the border differs from the number of commuters between the two countries. We assumed that the ratio of cars to commuters crossing the border is maintained in both directions:

(S2) $$\frac{C_{CA \to US}}{V_{CA \to US}} = \frac{C_{US \to CA}}{V_{US \to CA}}$$

| | |
|---|---|
| $C_{CA \to US}$ | Total daily commuters from Canada to US |
| $V_{CA \to US} = 5{,}546$ | Daily Canadian vehicles (cars) re-entering Canada after a one-day trip (daily average from Canadian frontier counts of 2015) |
| $C_{US \to CA} = 4{,}605$ | Total daily commuters form US to Canada (from 2011-2015 US census) |
| $V_{US \to CA} = 13{,}017$ | Daily US vehicles (cars) entering Canada for a one-day trip (daily average from Canadian frontier counts of 2015) |

Solving (S2) for $C_{CA \to US}$ we obtain:

(S3) $$C_{CA \to US} = V_{CA \to US} \frac{C_{US \to CA}}{V_{US \to CA}} \approx 1{,}962$$

The US commuters to Canada accounts for around 35% of the total US vehicles crossing the Canadian border daily. Therefore, we multiplied the number of Canadian cars crossing the US border daily by the same factor.

Finally, the total US commuters to Canada were distributed to the Canadian provinces/territories using formula (S1), where $C_{m^{B \to A}}$ are the Canadian commuters to the US from each province or territory and $C_{B \to A}$ is the total Canadian commuters to US.

***Supplementary Table 1**: 20 most abundant commuting flows across locations in the north American region*

| Residence Location | | Work Location | | Count |
|---|---|---|---|---|
| MX | Estado de México | MX | Distrito Federal (Mexico City) | 1,059,180 |
| US | New Jersey | US | New York | 416,871 |
| US | Maryland | US | District of Columbia (Washington DC) | 320,762 |
| CA | Quebec | CA | Ontario | 240,660 |
| US | Virginia | US | District of Columbia (Washington DC) | 228,039 |
| MX | Tlaxcala | MX | Puebla | 175,310 |
| US | New York | US | New Jersey | 127,032 |
| US | Maryland | US | Virginia | 125,267 |
| US | New Jersey | US | Pennsylvania | 125,247 |
| US | Pennsylvania | US | New Jersey | 124,808 |
| CA | Ontario | CA | Quebec | 104,140 |
| MX | Distrito Federal (Mexico City) | MX | Estado de México | 101,260 |
| US | Missouri | US | Kansas | 99,778 |
| US | Kansas | US | Missouri | 89,681 |
| US | New Hampshire | US | Massachusetts | 85,262 |
| US | Illinois | US | Missouri | 83,063 |
| US | Connecticut | US | New York | 73,539 |
| US | Washington | US | Oregon | 72,457 |
| US | Virginia | US | Maryland | 71,356 |
| US | South Carolina | US | North Carolina | 67,325 |

### Supplementary Note 2:

#### Metapopulation SEIR dynamical model

In this work we developed a metapopulation SEIR model to simulate COVID-19 transmission across the North American Region, encompassing Canadian provinces and territories, US states, and Mexican states. The model accounts for both documented and undocumented infections, with separate transmission rates defined for each. We assume no reinfection with SARS-CoV-2 and no vaccine effect on population susceptibility given their deployment on general population in the first months of 2021. Our model incorporates two forms of human mobility across the 96 locations: regular daily commuting and diffusive random movement. Commuters travel to their workplaces during the day, interacting with the local population, and return home in the evening, mixing with individuals in their residential areas. Besides commuting, a portion of the population in each location, proportional to inter-location commuters, travels for non-work purposes in random locations. As population dynamics differ between daytime and nighttime, we separately model COVID-19 transmission dynamics for these periods. This involves formulating transmission as a discrete Markov process and employing random sampling from Poisson distributions to capture stochastic model dynamics. The transmission dynamics are expressed through the following equations, the description of variables and parameters can be found in **Supplementary Table 2**.

**Daytime transmission**

(S4) $S_{ij}(t + dt_1) = S_{ij}(t) - Pois\left(\beta_i \frac{S_{ij}(t)\sum_k I_{ki}^r(t)}{N_i^d(t)} dt_1\right) - Pois\left(\mu\beta_i \frac{S_{ij}(t)\sum_k I_{ik}^u(t)}{N_i^d(t)} dt_1\right) +$

$Pois\left(\theta dt_1 \frac{N_{ij}-I_{ij}^r(t)}{N_i^d(t)} \sum_{k\neq i} \frac{\bar{N}_{ik}\sum_l S_{kl}(t)}{N_k^d(t)-\sum_l I_{lk}^r(t)}\right) - Pois\left(\theta dt_1 \frac{S_{ij}(t)}{N_i^d(t)-\sum_l I_{li}^r(t)} \sum_{k\neq i} \bar{N}_{ki}\right)$

(S5) $E_{ij}(t + dt_1) = E_{ij}(t) + Pois\left(\beta_i \frac{S_{ij}(t)\sum_k I_{ki}^r(t)}{N_i^d(t)} dt_1\right) + Pois\left(\mu\beta_i \frac{S_{ij}(t)\sum_k I_{ik}^u(t)}{N_i^d(t)} dt_1\right) - Pois\left(\frac{E_{ij}(t)}{Z} dt_1\right) +$

$Pois\left(\theta dt_1 \frac{N_{ij}-I_{ij}^r(t)}{N_i^d(t)} \sum_{k\neq i} \frac{\bar{N}_{ik}\sum_l E_{kl}(t)}{N_k^d(t)-\sum_l I_{lk}^r(t)}\right) - Pois\left(\theta dt_1 \frac{E_{ij}(t)}{N_i^d(t)-\sum_l I_{li}^r(t)} \sum_{k\neq i} \bar{N}_{ki}\right)$

(S6) $I_{ij}^r(t + dt_1) = I_{ij}^r(t) + Pois\left(\alpha \frac{E_{ij}(t)}{Z} dt_1\right) - Pois\left(\frac{I_{ij}^r(t)}{D} dt_1\right)$

(S7) $I_{ij}^u(t + dt_1) = I_{ij}^u(t) + Pois\left((1-\alpha) \frac{E_{ij}(t)}{Z} dt_1\right) - Pois\left(\frac{I_{ij}^u(t)}{D} dt_1\right) +$

$Pois\left(\theta dt_1 \frac{N_{ij}-I_{ij}^r(t)}{N_i^d(t)} \sum_{k\neq i} \frac{\bar{N}_{ik}\sum_l I_{kl}^u(t)}{N_k^d(t)-\sum_l I_{lk}^r(t)}\right) - Pois\left(\theta dt_1 \frac{I_{ij}^u(t)}{N_i^d(t)-\sum_l I_{li}^r(t)} \sum_{k\neq i} \bar{N}_{ki}\right)$

(S8) $N_i^d(t) = N_{ii} + \sum_{k\neq i} I_{ki}^r(t) + \sum_{k\neq i}(N_{ik} - I_{ik}^r(t))$

**Nighttime transmission**

(S9) $S_{ij}(t + 1) = S_{ij}(t + dt_1) - Pois\left(\beta_j \frac{S_{ij}(t+dt_1)\sum_k I_{kj}^r(t+dt_1)}{N_j^n} dt_2\right) -$

$Pois\left(\mu\beta_j \frac{S_{ij}(t+dt_1)\sum_k I_{kj}^u(t+dt_1)}{N_j^n} dt_2\right) + Pois\left(\theta dt_2 \frac{N_{ij}}{N_j^n} \sum_{k\neq j} \frac{\bar{N}_{jk}\sum_l S_{lk}(t+dt_1)}{N_k^n-\sum_l I_{lk}^r(t+dt_1)}\right) -$

$Pois\left(\theta dt_2 \frac{S_{ij}(t+dt_1)}{N_j^n-\sum_k I_{kj}^r(t+dt_1)} \sum_{k\neq j} \bar{N}_{kj}\right)$

(S10) $E_{ij}(t+1) = E_{ij}(t+dt_1) + Pois\left(\beta_j \frac{S_{ij}(t+dt_1)\sum_k I_{kj}^r(t+dt_1)}{N_j^n} dt_2\right) +$

$Pois\left(\mu\beta_j \frac{S_{ij}(t+dt_1)\sum_k I_{kj}^u(t+dt_1)}{N_j^n} dt_2\right) - Pois\left(\frac{E_{ij}(t+dt_1)}{Z} dt_2\right) + Pois\left(\theta dt_2 \frac{N_{ij}}{N_j^n} \sum_{k\neq j} \frac{\bar{N}_{jk}\sum_l E_{lk}(t+dt_1)}{N_k^n - \sum_l I_{lk}^r(t+dt_1)}\right) -$

$Pois\left(\theta dt_1 \frac{E_{ij}(t+dt_1)}{N_j^n - \sum_k I_{kj}^r(t+dt_1)} \sum_{k\neq j} \bar{N}_{kj}\right)$

(S11) $I_{ij}^r(t+1) = I_{ij}^r(t+dt_1) + Pois\left(\alpha \frac{E_{ij}(t+dt_1)}{Z} dt_2\right) - Pois\left(\frac{I_{ij}^r(t+dt_1)}{D} dt_2\right)$

(S12) $I_{ij}^u(t+1) = I_{ij}^u(t+dt_1) + Pois\left((1-\alpha)\frac{E_{ij}(t+dt_1)}{Z} dt_2\right) - Pois\left(\frac{I_{ij}^u(t+dt_1)}{D} dt_2\right) +$

$Pois\left(\theta dt_2 \frac{N_{ij}}{N_j^n} \sum_{k\neq j} \frac{\bar{N}_{jk}\sum_l I_{lk}^u(t+dt_1)}{N_k^n - \sum_l I_{lk}^r(t+dt_1)}\right) - Pois\left(\theta dt_2 \frac{I_{ij}^u(t+dt_1)}{N_j^n - \sum_k I_{kj}^r(t+dt_1)} \sum_{k\neq j} \bar{N}_{ki}\right)$

(S13) $N_i^n = \sum_k N_{ki}$

***Supplementary Table 2**: Description of state variables and parameters*

| | |
|---|---|
| $S_{ij}, E_{ij}, I_{ij}^r, I_{ij}^u, N_{ij}$ | Susceptible, exposed, reported infected, unreported infected, and total population in the subpopulations commuting from location *j* to location *i* (*i* ←*j*) |
| $\beta_i$ | Transmission rate of reported infections in state *i* |
| $\mu$ | Relative transmissibility of unreported infections |
| $Z$ | Average latency period from Infection to contagiousness |
| $D$ | Average duration of contagiousness |
| $\alpha$ | Fraction of documented infections |
| $\theta$ | Multiplicative factor adjusting random movements |
| $\bar{N}_{ij} = (N_{ij} + N_{ji})/2$ | Average number of commuters between state *i* and state *j* |
| $dt_1, dt_2$ | Daytime and nighttime duration $dt_1 + dt_2 = 1$ |
| $N_i^d, N_i^n$ | Daytime and nighttime population of state *i* |

We assume that the portion of the infectious population that is reported, $I_{ij}^r$, remains immobile and does not partake in human movement. We integrate Eqs. **S4-S13** using a Poisson process to represent the stochastic nature of the transmission process. During the daytime, $N_{ij}$ individuals residing in location $j$ commute to workplace $i$, interacting with the local population. At nighttime, these commuters return home and mix with other residents in location $j$. Additionally, individuals engage in random movement between locations for purposes other than work, circulating among subpopulations following a Markov process and contributing to population exchange across all locations.

**Daily work commuting**. For the daily work commuting, the daytime population in location $i$, $N_i^d(t) = N_{ii} + \sum_{k\neq i} I_{ki}^r(t) + \sum_{k\neq i}(N_{ik} - I_{ik}^r(t))$, is composed of individuals who both live and work in $i$, reported infected individuals who would otherwise commute to other locations $k\ (k \neq i)$, and individuals commuting to $i$ from other location $k\ (k \neq i)$ who are not reported infections. New infections of any subpopulation $N_{ij}$ occurs through contact with documented and with undocumented infectious individuals. Specifically, for each susceptible individual at time t $S_{ij}(t)$, the probability of encountering a documented infectious person is $\sum_k I_{ki}^r(t)/ N_i^d(t)$, where $\sum_k I_{ki}^r(t)$ is the number of documented infectious individuals in location $i$. These encounters create $\beta_i \frac{S_{ij}(t)\sum_k I_{ki}^r(t)}{N_i^d(t)} dt_1 + \mu\beta_i \frac{S_{ij}(t)\sum_k I_{ik}^u(t)}{N_i^d(t)} dt_1$ new infections during the daytime period $dt_1$, as indicated in **Eq. S4-S5**. This term accounts for the mixing of populations from different locations due to work commuting and represents intra-location transmission during the daytime in location $i$.

**Random movement.** In addition to people commuting for work, during daytime, $\theta dt_1 \bar{N}_{ik}$ people are drawn uniformly from population $k$ $(k \neq i)$ and are randomly distributed to the subpopulation of location $i$ (reported documented infectious individuals are not drawn as they stay home). This exchange exists for each pair of locations. First, we compute the number of susceptible individuals entering the subpopulation $S_{ij}(t)$. In the other locations $k$ $(k \neq i)$ the probability that a random visitor is susceptible is $\sum_l S_{kl}(t)/\,(N_k^d(t) - \sum_l I_{lk}^r(t))$ where $\sum_l S_{kl}(t)$ is the susceptible individuals that moved to location $k$ form any location $l$, while $N_k^d(t) - \sum_l I_{lk}^r(t)$ is the total population minus the reported infected in location $k$. Therefore, the total susceptible population entering location $i$ is $\theta dt_1 \bar{N}_{ik} \sum_{k\neq i} \sum_l S_{kl}(t)/\,(N_k^d(t) - \sum_l I_{lk}^r(t))$. These individuals are redistributed into the subpopulation of location $i$, here the people in the subpopulation $N_{ij}$ is $(N_{ij} - I_{ij}^r(t))/N_i^d(t)$. Finally, the susceptible individuals entering $S_{ij}(t)$ for the random movement can be computed as: $\theta dt_1 \frac{N_{ij} - I_{ij}^r(t)}{N_i^d(t)} \sum_{k\neq i} \frac{\bar{N}_{ik} \sum_l S_{kl}(t)}{N_k^d(t) - \sum_l I_{lk}^r(t)}$.

To compute the number of individuals leaving $S_{ij}(t)$ through the random movement process, we computed the number of individuals leaving location $i$: $\theta dt_1 \sum_{k\neq i} \bar{N}_{ki}$. Then, the computed the susceptible people coming from $N_{ij}$ is $S_{ij}(t)/(N_i^d(t) - \sum_l I_{li}^r(t))$. The resulting number of susceptible individuals leaving $S_{ij}(t)$ is thus $\theta dt_1 \frac{S_{ij}(t)}{N_i^d(t) - \sum_l I_{li}^r(t)} \sum_{k\neq i} \bar{N}_{ki}$.

The random movement (i.e. exchange of individuals) among other compartments can be computed similarly. **Eq. S6** does not have terms related to random movement: reported infectious individuals are assumed not to move from their subpopulation. Similar to daytime, **Eq. S9-S13** represent transmission during nighttime.

### Supplementary Note 3:

#### Ensemble Adjustment Kalman Filter (EAKF)

The ensemble adjustment Kalman filter (EAKF) was originally designed for weather forecasting and operates under the assumption that both the prior and the likelihood follow a normal Gaussian distribution. It deterministically transforms the prior distribution into a posterior using Bayes' rule. The EAKF represents the state-space distribution using an ensemble of system state vectors as samples. The assumption of Gaussian distributions for both the prior and likelihood enables characterization by their first two moments, the mean and variance. The ensemble members update occurs following Bayes' rule (posterior ∝ prior x likelihood). More specifically, the process involves the convolution of the two Gaussian distributions. The observed state variable (daily reported cases) are updated deterministically so that the higher moments of the prior distribution are preserved in the posterior. The *i*th ensemble member is updated at each timestep as:

(S14) $$o_{t,post}^{i}=\frac{\sigma_{t,obs}^{2}}{\sigma_{t,obs}^{2}+\sigma_{t,prior}^{2}}\bar{o}_{t,prior}+\frac{\sigma_{t,prior}^{2}}{\sigma_{t,obs}^{2}+\sigma_{t,prior}^{2}}y_t+\sqrt{\frac{\sigma_{t,obs}^{2}}{\sigma_{t,obs}^{2}+\sigma_{t,prior}^{2}}}(o_{t,prior}^{i}-\bar{o}_{t,prior})$$

| | |
|---|---|
| $o_{t,post}^{i}$ | Posterior of the observed variable |
| $o_{t,prior}^{i}$ | Prior of the observed variable for the *i*th ensemble member at time *t* |
| $\bar{o}_{t,prior}$ | Mean of the prior observed variable at time *t* |
| $\sigma_{t,obs}^{2}$ | Variance of the observed variable |
| $\sigma_{t,prior}^{2}$ | Variance of the prior observed variable |
| $y_t$ | Observation at time at time *t* |

The unobserved variables and parameters are also updated daily using their covariability with the observed variable. Their covariability can be computed directly from the ensemble. The *i*th ensemble member of an unobserved state variable or unobserved parameter is update at each timestep as:

(S15) $$x_{t,post}^{i}=x_{t,prior}^{i}+\frac{\sigma\left(\{x_{t,prior}\}_n,\{o_{t,prior}\}_n\right)}{\sigma_{t,prior}^{2}}(o_{t,post}^{i}-o_{t,prior}^{i})$$

| | |
|---|---|
| $x_{t,post}^{i}$ | Posterior of unobserved state variable or parameter for the *i*th ensemble member at time *t* |
| $x_{t,prior}^{i}$ | Prior of unobserved state variable or parameter for the *i*th ensemble member at time *t* |
| $\sigma\left(\{x_{t,prior}\}_n,\{o_{t,prior}\}_n\right)$ | Covariance between the priors of the of unobserved state variable or parameter and the observed state variable |

### Supplementary Note 4:

#### Exceptions to the national trends in the estimated parameters

The estimated values shown in **Figure 3-5** and **Supplementary Table 3** tend to follow very strong national trends, but there are some exceptions. Some Canadian territories and provinces, such as Northwest Territories, Nunavut, Prince Edward Island, and Yukon, exhibited relatively high transmission rates $\beta$ during the estimation period. However, it's important to note that their combined population represents less than 0.8% of the Canadian population and they reported extremely low daily case counts (i.e., <10). These factors contributed to increased stochasticity in the model for these locations. With such a low signal-to-noise ratio, the system may be less reliable at estimating parameter values.

In the US, notable exceptions to national trends are found in Alabama, North Dakota, and Ohio (see **Supplementary Figure 3**). Due to a backlog in reporting, Alabama added 4,007 cases data relative to test performed from October 2020 to January 2021 on March 15th 2021 (https://www.al.com/news/2021/03/alabama-adds-4556-covid-cases-4007-from-backlog-dating-back-to-2020.html), which is also the third timepoint selected to show the estimated values in this work. This event led the SEIR-EAKF system to adjust $\beta$ to high values (1.62). During the fall of 2020, North Dakota experienced a significant surge in cases. Following the outbreak, the estimated susceptible population dropped to approximately 30%, and by March 2021, $\beta$ value estimates became extremely broad. When transmission value estimates exceeded the bound values of 0.2 and 4, they were resampled across the current ensemble spread. When the spread is very large, the ensemble's average values will tend towards the midpoint between the upper and lower bounds (2.1). This could explain the exceptionally high estimate for North Dakota on March 31, 2021 (1.61). Ohio experienced a milder outbreak in spring 2020. The lower number of reported cases made it more challenging for the model to accurately estimate the force of infection ($\beta$) during this period. From January to March 2020, the estimated transmission values fluctuated unpredictably. This may explain the relatively high $\beta$ value for Ohio at the initial time point on June 6, 2020. As time progressed, the estimated values aligned with those estimated for other states across the country.

The hyperparameter $R_t$ (time-varying basic reproductive number) calculated by **eq. (1)**, is proportional to the values of $\beta$ and $\alpha$. For this reason, the $R_t$ of the locations with exceptionally high $\beta$ (described above) showed high basic reproductive number $R_t$.

***Supplementary Figure 1**: **a)** Total reported daily cases per country divided by the population. The dotted vertical lines indicates the three timepoints selected to show estimates in the main text. Source: [https://health.google.com/covid-19/open-data/data-sources](https://health.google.com/covid-19/open-data/data-sources) **b)** Percentage daily "change in workplace visitors" (i.e. individuals that commute to work) compared to baseline. The baseline is the median value, for the corresponding day of the week, during the 5-week period from January 3rd to February 6th 2020. The light curves represent each location, the thicker curves represent the average rate of the locations of each country. Source: [https://www.google.com/covid19/mobility/](https://www.google.com/covid19/mobility/)*

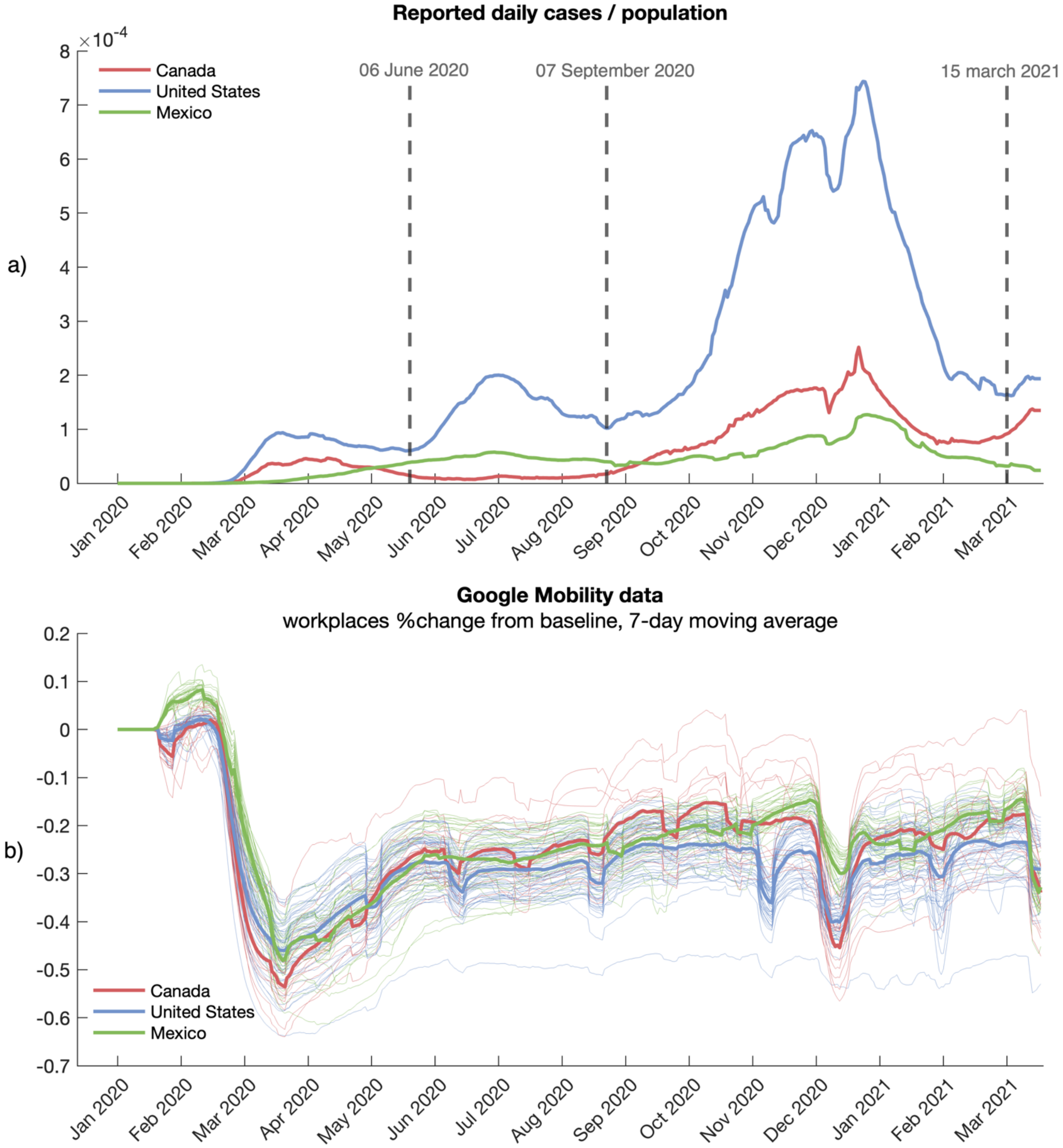

***Supplementary Figure 2**: Boxplot distributions of parameter estimates and synthetic true values. Each box represent the interquartile range (IQR) with the median line inside the box. Whiskers extend to the minimum and maximum values within 1.5 times the IQR; outliers are not shown.*

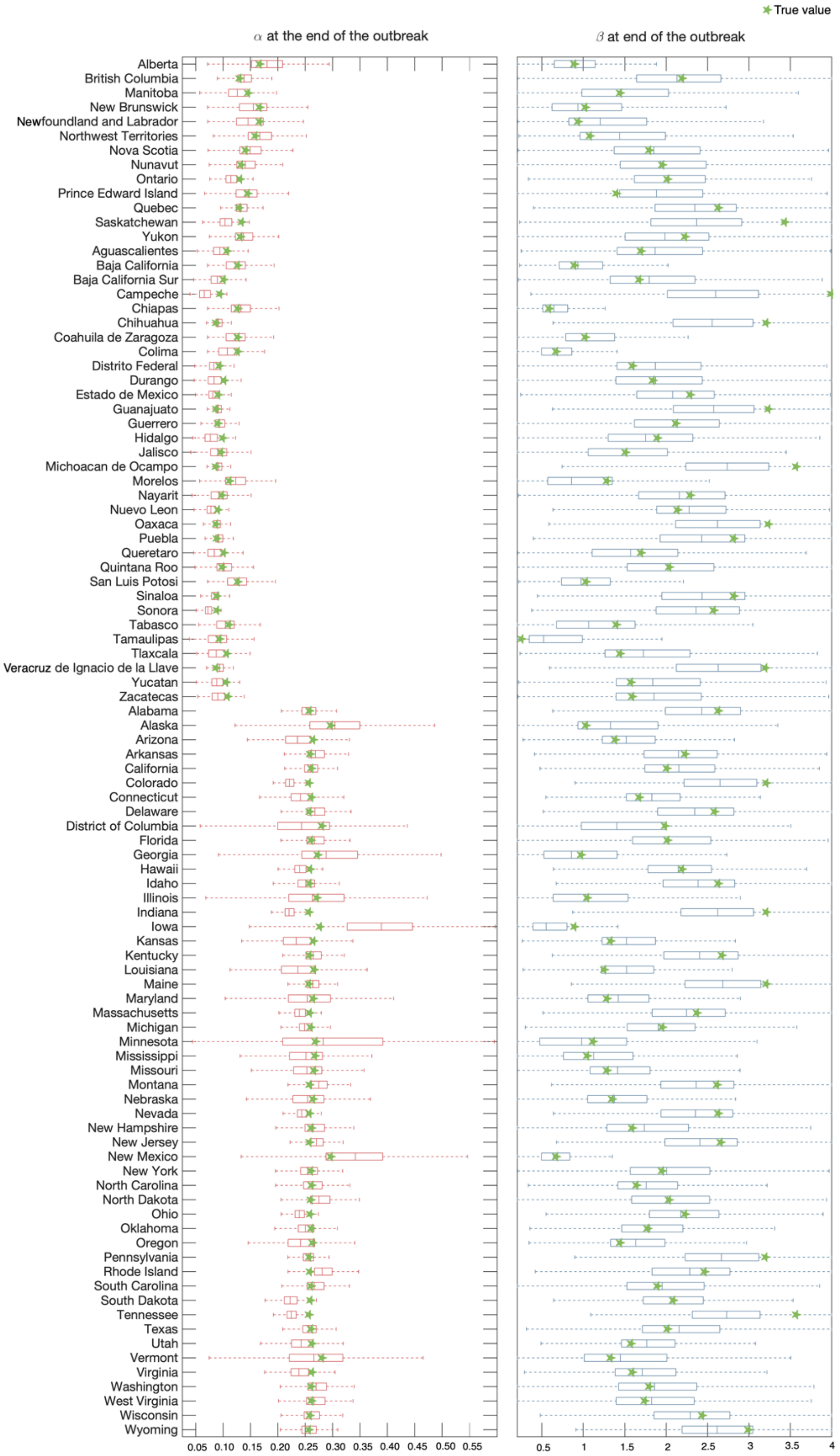

***Supplementary Figure 3**: Three US locations deviate from national trends in estimated parameters. Model fitting (i.e. estimated observed variable over the 7-day smoothed daily new reported cases, the model input), three state variables illustrating disease progression (the susceptible population, cumulative reported infectious and cumulative unreported infectious), and the parameters $\alpha$, $\beta$ and $R_t$ are shown for Alabama, North Dakota and Ohio. The color shaded areas represent the 95% credible interval from the 300-member ensemble. The dotted vertical lines indicate the three timepoints of reference (June 6, 2020; September 7, 2020; March 15, 2021)*

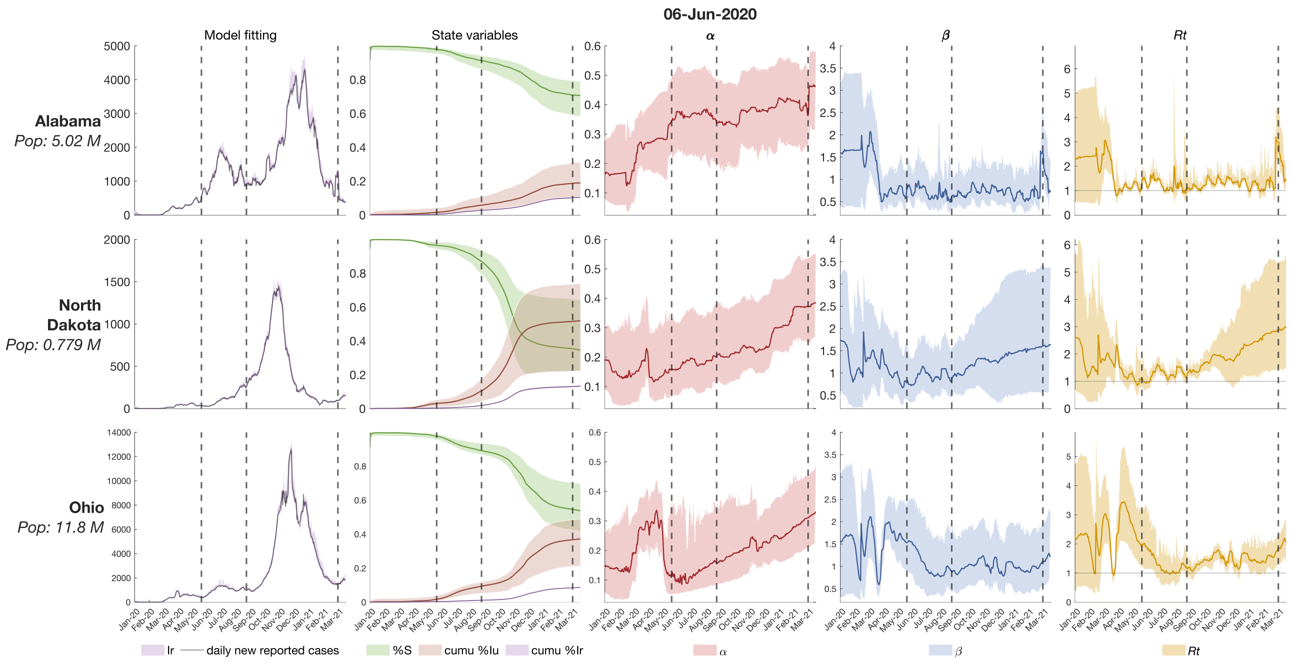

***Supplementary Table 3***: *Mean estimated values of the parameters (ascertainment rate $\alpha$ and transmission rate $\beta$) and hyperparameters (basic reproductive number $R_t$) for all the locations on the three selected timepoints. The intensity of cell colors in the table corresponds to their values: higher values are represented by more intense colors.*

| Location | | June 6th 2020 | | | September 7th 2020 | | | March 15th 2021 | | |
|---|---|---|---|---|---|---|---|---|---|---|
| | | $\alpha$ | $\beta$ | $R_t$ | $\alpha$ | $\beta$ | $R_t$ | $\alpha$ | $\beta$ | $R_t$ |
| CA | Alberta | 0.175 | 0.818 | 1.132 | 0.199 | 0.791 | 1.116 | 0.258 | 1.007 | 1.568 |
| CA | British Columbia | 0.192 | 0.690 | 0.959 | 0.192 | 0.853 | 1.229 | 0.259 | 0.724 | 1.117 |
| CA | Manitoba | 0.175 | 0.954 | 1.371 | 0.187 | 0.765 | 1.152 | 0.262 | 0.891 | 1.400 |
| CA | New Brunswick | 0.135 | 1.213 | 1.642 | 0.149 | 1.020 | 1.426 | 0.259 | 0.672 | 1.103 |
| CA | Newfoundland and Labrador | 0.162 | 1.091 | 1.635 | 0.162 | 1.232 | 1.775 | 0.293 | 1.083 | 1.869 |
| CA | Northwest Territories | 0.131 | 1.800 | 2.452 | 0.146 | 1.815 | 2.542 | 0.259 | 1.218 | 2.015 |
| CA | Nova Scotia | 0.127 | 0.715 | 1.012 | 0.136 | 0.941 | 1.298 | 0.257 | 0.628 | 1.047 |
| CA | Nunavut | 0.122 | 1.934 | 2.675 | 0.141 | 1.881 | 2.675 | 0.262 | 1.153 | 2.000 |
| CA | Ontario | 0.160 | 0.724 | 0.951 | 0.165 | 0.991 | 1.341 | 0.258 | 0.860 | 1.335 |
| CA | Prince Edward Island | 0.117 | 1.291 | 1.670 | 0.138 | 1.445 | 1.925 | 0.257 | 0.920 | 1.500 |
| CA | Quebec | 0.166 | 0.534 | 0.765 | 0.168 | 0.998 | 1.359 | 0.256 | 0.809 | 1.251 |
| CA | Saskatchewan | 0.192 | 0.755 | 1.136 | 0.293 | 0.919 | 1.537 | 0.317 | 0.645 | 1.077 |
| CA | Yukon | 0.125 | 1.366 | 1.821 | 0.143 | 1.421 | 1.919 | 0.263 | 1.120 | 1.836 |
| US | Alabama | 0.342 | 0.697 | 1.209 | 0.347 | 0.569 | 1.010 | 0.363 | 1.618 | 3.004 |
| US | Alaska | 0.282 | 1.093 | 1.764 | 0.358 | 0.651 | 1.143 | 0.424 | 0.676 | 1.268 |
| US | Arizona | 0.324 | 1.017 | 1.731 | 0.358 | 0.536 | 0.913 | 0.402 | 0.569 | 1.051 |
| US | Arkansas | 0.246 | 0.928 | 1.417 | 0.267 | 0.757 | 1.168 | 0.324 | 0.850 | 1.395 |
| US | California | 0.273 | 0.658 | 1.019 | 0.335 | 0.510 | 0.855 | 0.389 | 0.686 | 1.226 |
| US | Colorado | 0.335 | 0.495 | 0.826 | 0.365 | 0.579 | 1.022 | 0.407 | 0.573 | 1.076 |
| US | Connecticut | 0.191 | 0.598 | 0.818 | 0.238 | 0.890 | 1.347 | 0.348 | 1.046 | 1.823 |
| US | Delaware | 0.182 | 0.591 | 0.827 | 0.256 | 0.758 | 1.165 | 0.331 | 0.966 | 1.626 |
| US | District of Columbia (Washington DC) | 0.320 | 0.564 | 0.932 | 0.357 | 0.628 | 1.099 | 0.451 | 0.646 | 1.235 |
| US | Florida | 0.304 | 0.814 | 1.341 | 0.358 | 0.582 | 1.001 | 0.329 | 0.857 | 1.439 |
| US | Georgia | 0.281 | 0.741 | 1.170 | 0.286 | 0.630 | 0.985 | 0.304 | 1.104 | 1.832 |
| US | Hawaii | 0.252 | 0.864 | 1.356 | 0.371 | 0.499 | 0.895 | 0.437 | 0.654 | 1.268 |
| US | Idaho | 0.310 | 0.669 | 1.085 | 0.365 | 0.573 | 1.019 | 0.384 | 0.786 | 1.412 |
| US | Illinois | 0.255 | 0.509 | 0.760 | 0.308 | 0.703 | 1.155 | 0.347 | 0.917 | 1.585 |
| US | Indiana | 0.289 | 0.573 | 0.900 | 0.321 | 0.593 | 0.987 | 0.367 | 0.842 | 1.478 |
| US | Iowa | 0.220 | 0.766 | 1.126 | 0.290 | 0.628 | 1.009 | 0.402 | 0.843 | 1.552 |
| US | Kansas | 0.089 | 0.773 | 0.926 | 0.169 | 0.746 | 0.985 | 0.309 | 1.019 | 1.662 |
| US | Kentucky | 0.175 | 0.752 | 1.025 | 0.229 | 0.663 | 0.958 | 0.322 | 0.908 | 1.501 |
| US | Louisiana | 0.144 | 0.854 | 1.096 | 0.232 | 0.792 | 1.171 | 0.342 | 0.944 | 1.608 |
| US | Maine | 0.239 | 0.597 | 0.896 | 0.346 | 0.599 | 1.046 | 0.372 | 0.746 | 1.339 |
| US | Maryland | 0.203 | 0.607 | 0.840 | 0.267 | 0.785 | 1.228 | 0.340 | 0.865 | 1.489 |
| US | Massachusetts | 0.218 | 0.535 | 0.789 | 0.248 | 0.698 | 1.060 | 0.342 | 0.939 | 1.613 |
| US | Michigan | 0.289 | 0.793 | 1.478 | 0.315 | 0.769 | 1.301 | 0.337 | 1.092 | 1.885 |
| US | Minnesota | 0.314 | 0.487 | 0.801 | 0.359 | 0.527 | 0.920 | 0.394 | 0.839 | 1.544 |
| US | Mississippi | 0.341 | 0.720 | 1.283 | 0.327 | 0.560 | 0.924 | 0.385 | 0.840 | 1.499 |
| US | Missouri | 0.276 | 0.658 | 1.023 | 0.371 | 0.612 | 1.085 | 0.314 | 0.926 | 1.611 |
| US | Montana | 0.238 | 0.915 | 1.405 | 0.308 | 0.599 | 0.978 | 0.400 | 0.968 | 1.813 |
| US | Nebraska | 0.301 | 0.502 | 0.844 | 0.373 | 0.614 | 1.073 | 0.422 | 0.693 | 1.294 |

| Location | | June 6th 2020 | | | September 7th 2020 | | | March 15th 2021 | | |
|---|---|---|---|---|---|---|---|---|---|---|
| | | $\alpha$ | $\beta$ | $R_t$ | $\alpha$ | $\beta$ | $R_t$ | $\alpha$ | $\beta$ | $R_t$ |
| US | Nevada | 0.295 | 0.868 | 1.432 | 0.365 | 0.499 | 0.873 | 0.446 | 0.560 | 1.107 |
| US | New Hampshire | 0.150 | 0.639 | 0.832 | 0.219 | 0.926 | 1.372 | 0.389 | 0.788 | 1.454 |
| US | New Jersey | 0.277 | 0.549 | 0.904 | 0.352 | 0.684 | 1.182 | 0.357 | 0.915 | 1.601 |
| US | New Mexico | 0.199 | 0.928 | 1.315 | 0.278 | 0.562 | 0.908 | 0.363 | 0.770 | 1.355 |
| US | New York | 0.153 | 0.663 | 0.839 | 0.217 | 0.819 | 1.188 | 0.330 | 0.926 | 1.553 |
| US | North Carolina | 0.253 | 0.806 | 1.220 | 0.280 | 0.545 | 0.860 | 0.324 | 0.906 | 1.523 |
| US | North Dakota | 0.157 | 0.809 | 1.061 | 0.201 | 0.803 | 1.137 | 0.372 | 1.609 | 2.885 |
| US | Ohio | 0.109 | 1.531 | 1.928 | 0.161 | 0.914 | 1.217 | 0.310 | 1.107 | 1.825 |
| US | Oklahoma | 0.314 | 0.690 | 1.150 | 0.361 | 0.673 | 1.182 | 0.361 | 0.729 | 1.271 |
| US | Oregon | 0.290 | 0.976 | 1.602 | 0.312 | 0.576 | 0.950 | 0.366 | 0.596 | 1.047 |
| US | Pennsylvania | 0.202 | 0.583 | 0.809 | 0.267 | 0.741 | 1.143 | 0.363 | 0.872 | 1.544 |
| US | Rhode Island | 0.273 | 0.536 | 0.838 | 0.387 | 0.650 | 1.183 | 0.428 | 0.797 | 1.511 |
| US | South Carolina | 0.211 | 0.979 | 1.405 | 0.272 | 0.674 | 1.041 | 0.327 | 0.913 | 1.531 |
| US | South Dakota | 0.371 | 0.593 | 1.054 | 0.416 | 0.557 | 1.020 | 0.384 | 0.902 | 1.643 |
| US | Tennessee | 0.269 | 0.689 | 1.059 | 0.289 | 0.684 | 1.090 | 0.335 | 1.221 | 2.089 |
| US | Texas | 0.334 | 0.687 | 1.155 | 0.382 | 0.502 | 0.899 | 0.458 | 0.600 | 1.151 |
| US | Utah | 0.249 | 0.896 | 1.349 | 0.287 | 0.717 | 1.146 | 0.369 | 0.874 | 1.536 |
| US | Vermont | 0.290 | 1.038 | 1.732 | 0.354 | 0.607 | 1.157 | 0.464 | 0.567 | 1.123 |
| US | Virginia | 0.274 | 0.522 | 0.814 | 0.286 | 0.685 | 1.081 | 0.390 | 0.775 | 1.414 |
| US | Washington | 0.223 | 0.809 | 1.181 | 0.247 | 0.649 | 0.974 | 0.316 | 0.749 | 1.242 |
| US | West Virginia | 0.376 | 0.804 | 1.658 | 0.401 | 0.603 | 1.097 | 0.429 | 0.754 | 1.437 |
| US | Wisconsin | 0.306 | 0.524 | 0.847 | 0.307 | 0.821 | 1.369 | 0.366 | 0.966 | 1.727 |
| US | Wyoming | 0.222 | 0.712 | 1.051 | 0.308 | 0.719 | 1.210 | 0.341 | 0.818 | 1.407 |
| MX | Aguascalientes | 0.100 | 1.049 | 1.247 | 0.108 | 0.716 | 0.865 | 0.126 | 0.807 | 0.999 |
| MX | Baja California | 0.104 | 0.976 | 1.173 | 0.113 | 0.953 | 1.154 | 0.160 | 0.719 | 0.955 |
| MX | Baja California Sur | 0.097 | 1.180 | 1.402 | 0.105 | 0.993 | 1.196 | 0.127 | 1.155 | 1.444 |
| MX | Campeche | 0.078 | 1.242 | 1.451 | 0.116 | 0.672 | 0.824 | 0.123 | 0.854 | 1.060 |
| MX | Chiapas | 0.086 | 0.852 | 0.970 | 0.109 | 0.836 | 1.028 | 0.146 | 0.686 | 0.886 |
| MX | Chihuahua | 0.076 | 0.858 | 0.971 | 0.085 | 0.855 | 0.982 | 0.114 | 0.953 | 1.162 |
| MX | Coahuila | 0.089 | 1.141 | 1.340 | 0.098 | 0.779 | 0.919 | 0.136 | 0.720 | 0.914 |
| MX | Colima | 0.073 | 1.202 | 1.367 | 0.104 | 0.740 | 0.876 | 0.128 | 0.833 | 1.040 |
| MX | Distrito Federal (Mexico City) | 0.113 | 0.887 | 1.074 | 0.115 | 0.966 | 1.176 | 0.134 | 1.594 | 2.007 |
| MX | Durango | 0.074 | 1.361 | 1.557 | 0.094 | 0.876 | 1.019 | 0.123 | 0.977 | 1.209 |
| MX | Estado de México | 0.093 | 0.963 | 1.136 | 0.100 | 0.889 | 1.059 | 0.120 | 0.729 | 0.900 |
| MX | Guanajuato | 0.104 | 1.124 | 1.344 | 0.115 | 0.838 | 1.021 | 0.134 | 0.878 | 1.120 |
| MX | Guerrero | 0.104 | 1.032 | 1.217 | 0.123 | 1.029 | 1.294 | 0.150 | 0.755 | 0.980 |
| MX | Hidalgo | 0.109 | 0.866 | 1.049 | 0.115 | 0.746 | 0.913 | 0.140 | 0.725 | 0.931 |
| MX | Jalisco | 0.105 | 1.179 | 1.412 | 0.105 | 0.905 | 1.098 | 0.143 | 0.732 | 0.946 |
| MX | Michoacán | 0.105 | 1.017 | 1.226 | 0.108 | 0.846 | 1.021 | 0.119 | 0.841 | 1.042 |
| MX | Morelos | 0.095 | 0.885 | 1.041 | 0.102 | 0.787 | 0.938 | 0.138 | 0.738 | 0.916 |
| MX | Nayarit | 0.078 | 1.100 | 1.252 | 0.087 | 0.872 | 1.018 | 0.119 | 0.765 | 0.936 |
| MX | Nuevo León | 0.118 | 1.219 | 1.527 | 0.144 | 0.821 | 1.052 | 0.154 | 0.667 | 0.867 |
| MX | Oaxaca | 0.096 | 1.052 | 1.239 | 0.103 | 0.984 | 1.192 | 0.116 | 0.777 | 0.947 |
| MX | Puebla | 0.095 | 1.115 | 1.336 | 0.109 | 0.772 | 0.935 | 0.118 | 0.822 | 1.015 |
| MX | Querétaro | 0.109 | 0.936 | 1.138 | 0.114 | 0.742 | 0.901 | 0.136 | 0.856 | 1.066 |
| MX | Quintana Roo | 0.076 | 0.939 | 1.065 | 0.089 | 0.805 | 0.925 | 0.135 | 0.923 | 1.171 |
| MX | San Luis Potosí | 0.108 | 1.325 | 1.630 | 0.130 | 0.756 | 0.945 | 0.155 | 0.810 | 1.067 |

| Location | | June 6th 2020 | | | September 7th 2020 | | | March 15th 2021 | | |
|---|---|---|---|---|---|---|---|---|---|---|
| | | $\alpha$ | $\beta$ | $R_t$ | $\alpha$ | $\beta$ | $R_t$ | $\alpha$ | $\beta$ | $R_t$ |
| MX | Sinaloa | 0.071 | 1.070 | 1.208 | 0.083 | 0.899 | 1.029 | 0.105 | 0.949 | 1.134 |
| MX | Sonora | 0.095 | 0.980 | 1.160 | 0.110 | 0.826 | 1.007 | 0.141 | 0.964 | 1.223 |
| MX | Tabasco | 0.110 | 1.126 | 1.397 | 0.123 | 0.756 | 0.926 | 0.142 | 0.880 | 1.114 |
| MX | Tamaulipas | 0.094 | 1.092 | 1.298 | 0.117 | 0.931 | 1.142 | 0.164 | 0.781 | 1.030 |
| MX | Tlaxcala | 0.078 | 0.995 | 1.142 | 0.098 | 0.805 | 0.948 | 0.134 | 0.904 | 1.156 |
| MX | Veracruz | 0.084 | 0.931 | 1.071 | 0.091 | 0.844 | 0.982 | 0.119 | 0.746 | 0.916 |
| MX | Yucatán | 0.086 | 0.999 | 1.184 | 0.114 | 0.870 | 1.066 | 0.157 | 0.811 | 1.058 |
| MX | Zacatecas | 0.076 | 1.217 | 1.403 | 0.109 | 0.823 | 0.993 | 0.141 | 0.804 | 1.018 |
